# Hypoferremia reduces long-term risk of major adverse cardiovascular events after STEMI by averting the myocardial reactive iron storm

**DOI:** 10.1101/2025.01.13.25320244

**Authors:** M Vera-Aviles, S.N Kabir, M.D Christodoulou, G Mohammad, M Shanmuganathan, R Kotronias, J.G.F. Cleland, OxAMI investigators, KM Channon, S Lakhal-Littleton

**Author notes:** Corresponding author Samira Lakhal-Littleton. Department of Physiology, Anatomy & Genetics, University of Oxford, Sherrington Building, Parks Road, Oxford OX1 3PT, United Kingdom.

## Abstract

**Background and aims:** Iron deficiency (ID) is common in patients with acute STEMI. ID has previously been associated with either adverse or favourable effects, depending on the definition of ID, sampling timepoint, outcome measures and follow-up duration. This study systematically addresses the impact of ID on long-term outcomes and explores the underlying mechanisms.

**Methods:** Patients with acute STEMI (n=167) were followed for 4.5 years for major adverse cardiovascular events (MACE), including new HF diagnosis, recurrent MI or cardiac death. Iron markers were sampled at presentation and later timepoints. Myocardial injury was assessed by CMR at 2 days and 6 months. Mechanisms underlying clinical findings were evaluated in a mouse model of MI.

**Results:** ID, by any definition, was common in acute STEMI patients at presentation. Hypoferremia (Tsat<20% or iron<13uM) but not hypoferritinemia (ferritin<100ug/L) predicted lower risk of MACE. Hypoferremia predicted lower troponin, acute myocardial T1, LVESV and LVEDV at 2 days, but lower myocardial salvage at 6 months. Iron status sampled from 6 hours after presentation was no longer associated with MACE.

In mice, MI rapidly triggered a myocardial reactive iron storm, early LV remodelling, and eventually HF. These effects were averted by iron restriction.

**Conclusions:** This study reveals that hypoferremia in acute STEMI exerts both favourable and adverse effects, that nonetheless translate into better long-term clinical outcomes. This research reconciles previous seemingly conflicting reports. It also highlights a potential stepwise approach of acute iron chelation to reduce myocardial injury, followed days later by iron supplementation to promote myocardial salvage.

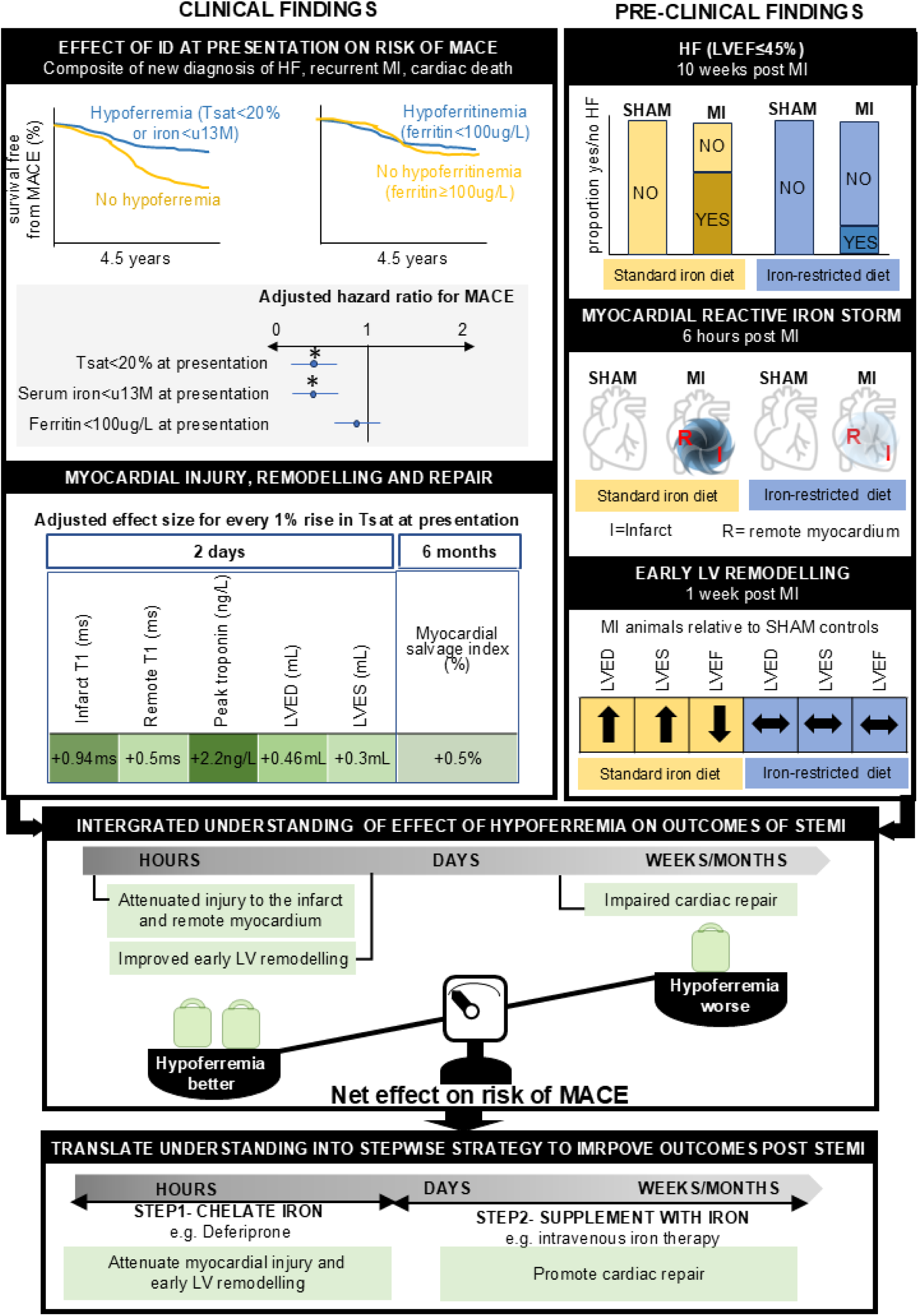

## Introduction

Despite major advances in the treatment of patients with acute ST elevation myocardial infarction (STEMI), a substantial proportion go on to develop heart failure (HF) or experience another major adverse cardiovascular event (MACE) (1,2). There is a pertinent need to identify and address the risk factors for MACE after STEMI. Iron deficiency (ID) is clinically defined as the presence of hypoferremia (i.e. low transferrin saturation Tsat, or low serum iron), or hypoferritinemia (i.e. low serum ferritin) (3). ID, by any definition, is prevalent in STEMI patients (4–8). Whether and how it affects outcomes remain unresolved. Past studies have yielded seemingly conflicting results. On the one hand, the presence of ID, defined according to the current European Society of Cardiology’s definition (ferritin<100ug/L, or Tsat<20% with ferritin 100-299ug/L) on the morning after STEMI, was associated with larger infarcts and more extensive microvascular obstruction (MVO) in the days after STEMI, and with higher frequency of adverse left ventricular (LV) remodelling at 6 months (4). Patients with serum iron below 7.8uM, before initiation of percutaneous coronary intervention (PCI), were found to have higher in-hospital mortality rates (5). Serum iron≤13uM measured on the day after hospital admission was found to predict higher all-cause mortality over a follow-up period of 3.4 years (6). On the other hand, ID, defined as Tsat< 20%, or serum ferritin<100 μg/L at hospital admission, was found to be associated with a lower incidence of a composite outcome of in-hospital mortality and Killip class≥3 (7). STEMI and Non-STEMI patients in the upper quartile of serum iron measured within 72 hours of onset of symptoms, had higher all-cause mortality at 10-month follow-up (8). These studies differed from one another in terms the definition of ID, the timing at which iron markers were sampled, the outcome measures being assessed, and the length of follow-up. These differences likely contributed to the divergence of results. Indeed, the different definitions of ID denote distinct phenomena. While hypoferremia is a reduction in the immediately accessible and reactive pool of iron, hyperferritinemia reflects a reduction in central reserves of non-reactive iron, which are not immediately accessible to tissues (9). Additionally, sampling in the days post STEMI may not reflect a patient’s baseline iron status, because STEMI itself is known to trigger rapid and transient changes in serum iron markers (10–12). Furthermore, different outcome measures and follow-up durations capture distinct processes in which iron is known to play either a favourable or an adverse role. For instance, iron is a catalyst for the initial cardiomyocyte death, and an exacerbating factor in subsequent ischemia-reperfusion injury (IRI) (10, 13–15). Conversely, iron is important for immune function, and oxidative phosphorylation that promote cardiac repair and contractility (16–18).

We sought to systematically address the question of whether a patient’s iron status affects outcomes after acute STEMI. We performed a retrospective longitudinal study in 167 patients admitted to hospital with acute STEMI. We evaluated long-term clinical outcomes over a median follow-up period of 4.5 years, by assessing the incidence of MACE, a composite outcome of new diagnosis of HF, cardiac death, or recurrent MI. First, to minimise any confounding effects of changes in iron markers secondary to STEMI, we inferred a patient’s iron status from the acute blood sample collected before emergency PCI. Second, we applied multiple definitions to determine the prevalence of ID, and to evaluate which definitions, if any, were associated with risk of MACE. Third, recognising that ID likely affected several processes, we examined the relationship between iron markers and multiple outcome measures of myocardial injury, remodelling and repair collected at 2 days and at 6 months post STEMI. Furthermore, we repeated all the above analyses using iron markers sampled in the hours and days post PCI, to determine if the sampling timepoint altered the results of analysis. Finally, we used a pre-clinical model of MI to validate and further extend the mechanistic insights derived from the clinical study.

## METHODS

### Study population

This was a retrospective sub-study within the Oxford Acute Myocardial Infarction (OxAMI) study. It included patients with STEMI admitted to Oxford Heart Centre for primary percutaneous coronary intervention PPCI between 2012 and 2018 and who were prospectively enrolled in OxAMI. The OxAMI study protocol was approved by the local ethics committee (REC: 11/SC/0397). Details of the OXAMI study protocol, inclusion and exclusion criteria were as published elsewhere (19, 20).

### Blood sampling and iron biomarkers

Iron markers were measured in serum extracted from blood collected at 6 sampling timepoints; at presentation (before PCI), immediately post PCI, 6 hours post PCI, 24 hours post PCI, 48 hours post PCI and at 6 month after discharge. Blood was collected from the aortic root before PCI and immediately post PCI, and from venous circulation at subsequent timepoints. Serum ferritin, transferrin and iron concentrations were determined using the ABX-Pentra C400 system (Horiba) as in our previous work (21).

### CMR and angiographic data

Acute CMR was performed within 48 hours of admission while follow-up CMR was performed at 6 months. Scans were performed on 3.0-T CMR scanners (either MAGNETOM Tim Trio or MAGNETOM Verio, Siemens Healthcare) as described previously (22). CMR images were analysed using the CVI42 software (Circle Cardiovascular Imaging Inc) and MC-ROI, a dedicated in-house software (programmed in IDL, version 8.7, L3Harris Geospatial) (20, 22). Parameters calculated from this analysis were Infarct size, the presence of microvascular obstruction (MVO), presence of intramyocardial haemorrhage (IMH), the size of the area at risk (AAR), and the relaxometry parameter T1, in the infarct and non-infarct. Thrombolysis In Myocardial Infarction (TIMI) flow pre and post PCI and myocardial blush grade (MBG) were derived as described previously (23).

### MACE

MACE was defined as a composite endpoint consisting of cardiac death, recurrent MI, or new diagnosis of HF. HF was defined as the new occurrence of symptomatic fluid overload for which diuretic agents were administered in patients with left ventricular ejection fraction (LVEF) <50% and/or raised levels of natriuretic peptides. Events before the acute CMR scan were excluded from analysis. Clinical outcomes data were collected from patients at their research visits, annual telephone calls, from electronic patient records and general practitioners.

### Mice

All animal procedures were compliant with the UK Home Office Mice (Scientific Procedures) Act 1986 (licence# P84F13B1B) and approved by the University of Oxford Medical Sciences Division Ethical Review Committee. B6;FVB mice were used, and both males and females included in the study cohort. To reflect the sex distribution of STEMI patients, the ratio of males to females in the mouse cohort was 4 to 1. Mice were randomly assigned to receive either a standard diet containing 200 ppm iron or an iron-restricted diet containing 5 ppm iron (Teklad TD.99397). Mice from each dietary group were randomised to two surgical interventions; MI or SHAM surgery.

### Procedures in mice

Details of MI surgery, cardiac imaging (Cine MRI) and reactive iron imaging are as described in previous work (21) and in supplemental methods.

## STATISTICS

All clinical data are reported as mean or median (IQ1-IQ3). Preclinical data are described as mean±standard error of the mean. Significance was set at p<0.05.

### Software and packages

All analysis was conducted using R (R version 4.4.1 (2024-06-14)), RStudio (2024.09.0+375), and a selection of packages, most importantly: tidyverse, MASS, lmtest, ggfortify, marginaleffects, compareGroups, betareg, syrvival, and survminer,

### Data pre-processing

Extensive integrity checks were performed using data visualisation, examination of extreme values and summary statistics. For MACE outcomes, data were filtered to 2700 days or less. Tsat at presentation was dichotomised at 20% and demographic summary statistics were calculated using median and IQR. The two groups were compared non-parametrically. Full cohort summary statistics were also calculated.

### Survival analysis

Events for MACE were first investigated using log rank test and Kaplan Meier survival curves for a variety of markers dichotomised at each sampling timepoints. Regression analysis was performed using Cox proportional hazards for each of the variables after ensuring no violations of proportionality. Although initially a mixed model approach was attempted to include all timepoints in a single analysis, the models did not converge, and the analysis was simplified to examine each timepoint separately. At a first instance, all relevant control variables were included, and the resulting models were examined using diagnostic plots and tests. To avoid overfitting, the full models were subsequently simplified using a manual step by step simplification informed by diagnostics, likelihood ratio testing and AIC. The simplified models are outlined in supplemental table 1. The most parsimonious model for each marker/timepoint combination was illustrated in a forest plot, and manual forest plots of all the markers by timepoint were created for ease of comparison of marker effect on MACE. To ensure that the results were not an artefact of variable dichotomisation, all analysis was re-run on the continuous marker estimates, with no differences observed.

### Regression analysis

Each acute or 6-month CMR outcome variable was examined for its distribution. Percentages were first analysed using a linear model and if diagnostics suggested a model misspecification, they were re-analysed using beta regression. Percentages that were at 0 or 100% were transformed as suggested by Smithson and Verkuilen (24). Binary variables were analysed using logistic regression. For ease of interpretation, odds ratios from beta and logistic regressions were transformed to average slopes which are the partial derivatives of the regression equation with respect to marker of interest. All other variables used a linear model, after checking assumptions through diagnostic plots and variance inflation factor for collinearity. As with the Cox regressions, to avoid overfitting, manual model simplification was conducted (supplemental table 1). To illustrate relative effect of Tsat on acute and 6-month CMR outcomes, the slope estimates (including average slope where relevant) where transformed on a −1 to +1 scale using min max normalisation. The visualisation is presented as a heatmap (figures 4 and 5).

### Preclinical data

In mice, multiple group comparisons were drawn by two-way ANOVA followed by Sidak’s post hoc test. Pairwise comparison of incidence of HF was drawn using two-tailed Fisher’s exact test.

## RESULTS

### Baseline characteristics and prevalence of ID

From the larger cohort of patients with acute STEMI who were enrolled in the OxAMI study, the present study identified 167 patients who underwent CMR before discharge (median 2 days post admission) and for whom sufficient serum volume was available from sampling timepoints before hospital discharge (Figure 1). In this cohort of 167 patients, the median age was 61.0 years (IQR: [54.0; 68.0]), 85% were male and 5.4% had a history of MI. The median pain-to-PCI time was 189 [125; 298] minutes and median peak Troponin was 49.4 [15.8; 90.6] ng/L. Table 1 summarises the levels of iron markers at presentation, baseline demographics, risk factors, clinical and angiographic characteristics as well as medications before STEMI, at discharge and at 6-month follow-up. Data are shown for the entire cohort, and stratified into two sub-groups according to Tsat<20% or Tsat≥20% at presentation.

**Figure 1.**
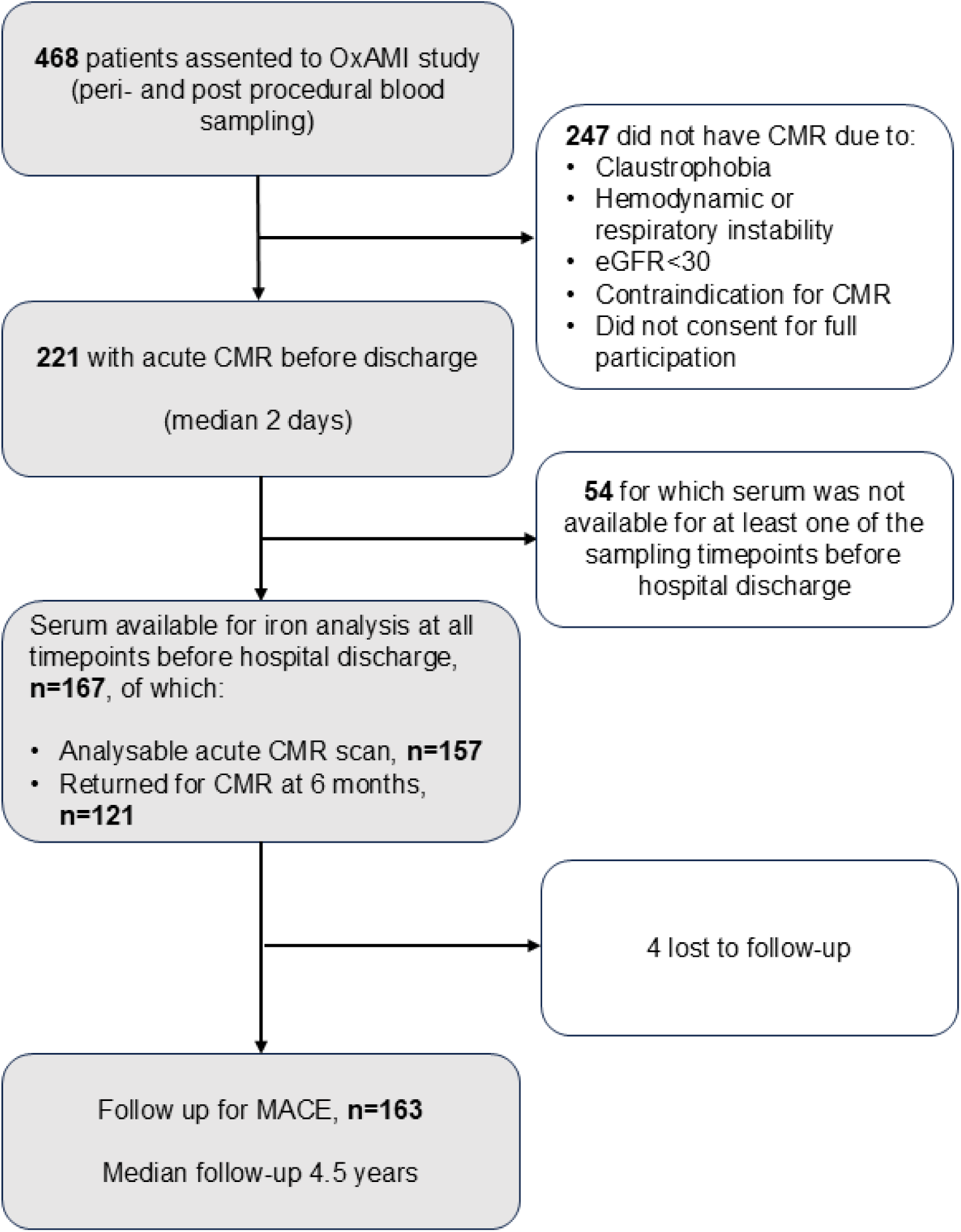
Study Profile

**Table 1:** Baseline characteristics, iron status in the entire cohort and dichotomised according to dichotomised Tsat at presentation.

|  | **All Cohort**<br>**(N=167)** | **Baseline Tsat<20%**<br>**(n=86, 54.09%)** | **Baseline Tsat≥20%**<br>**(n=73, 45.91%)** | **P value** |
| --- | --- | --- | --- | --- |
| **Iron status at admission** |  |  |  |  |
| **Tsat (%), median [IQR]** | 17.5<br>[11.1;34.7] | 11.5<br>[7.79;14.5] | 37.7<br>[28.5;49.0] | **<0.001** |
| **Ferritin (µg/L), median [IQR]** | 110<br>[64.8; 193] | 97.0<br>[55.4; 166] | 126<br>[71.2; 251] | **0.012** |
| **Ferritin<100µg/L , n (%)** | 75 (45.5%) | 45 (52.3%) | 27 (37.0%) | 0.076 |
| **Serum iron (µM), median [IQR]** | 11.4<br>[7.22; 16.7] | 9.06<br>[6.11; 12.1] | 15.8<br>[11.4; 20.2] | <0.001 |
| **Serum iron <13uM, n (%)** | 99 (60.0%) | 69 (80.2%) | 24 (32.9%) | **<0.001** |
| **Serum transferrin (g/L) median [IQR]** | 2.08<br>[1.64; 3.32] | 3.14<br>[2.24; 4.46] | 1.68 [1.44; 1.96] | **<0.001** |
| **ESC definition of ID** | 112 (69.1%) | 82 (95.3%) | 27 (37.0%) | **<0.001** |
| **(Ferritin<100µg/L or 100–299 µg/L with TSAT < 20%), n (%)** |  |  |  |  |
| **Demographics and risk factors** |  |  |  |  |
| **Gender, male, n (%)** | 141 (84.9%) | 70 (82.4%) | 65 (89.0%) | 0.336 |
| **Age, median [IQR]** | 61.0 [54.0; 68.0] | 61.0 [54.0; 68.0] | 61.0 [52.0; 67.0] | 0.633 |
| **BMI, median [IQR]** | 27.2 [24.6; 29.9] | 26.6 [24.2; 29.8] | 28.1 [25.7; 30.1] | 0.224 |
| **Smoking history, n (%)** | 113 (68.1%) | 62 (72.9%) | 44 (60.3%) | 0.129 |
| **Hypertension, n (%)** | 74 (44.3%) | 42 (48.8%) | 30 (41.1%) | 0.414 |
| **Diabetes, n (%)** | 35 (21.0%) | 14 (16.3%) | 19 (26.0%) | 0.189 |
| **Hypercholesterolemia, n (%)** | 59 (35.3%) | 29 (33.7%) | 28 (38.4%) | 0.659 |
| **Family history of CAD, n (%)** | 82 (49.4%) | 44 (51.8%) | 33 (45.2%) | 0.507 |
| **Previous MI, n (%)** | 9 (5.39%) | 5 (5.81%) | 4 (5.48%) | 1.000 |
| **Previous PCI, n (%)** | 6 (3.66%) | 4 (4.76%) | 2 (2.78%) | 0.831 |
| **COPD or Asthma, n (%)** | 18 (10.8%) | 6 (6.98%) | 10 (13.7%) | 0.254 |
| **Medications before STEMI** |  |  |  |  |
| **ACEI or ARB, n (%)** | 32 (19.2%) | 16 (18.6%) | 15 (20.5%) | 0.914 |
| **Beta blocker, n (%)** | 17 (10.2%) | 10 (11.6%) | 5 (6.85%) | 0.450 |
| **Statin, n (%)** | 34 (20.4%) | 19 (22.1%) | 13 (17.8%) | 0.636 |
| **Clinical and angiographic characteristics** |  |  |  |  |
| **BP systolic on admission mmHg, median [IQR]** | 128 [111; 150] | 126 [111; 149] | 128 [111; 145] | 0.952 |
| **HR on admission beats/min, median [IQR]** | 80 [65; 90] | 79.0 [64.0; 88.2] | 80.0 [70.0; 90.0] | 0.228 |
| **Pain to PCI time (min), mean [IQR]** | 189 [125; 298] | 176 [120; 317] | 189 [129; 258] | 0.981 |
| **Late presenter (>6h), n (%)** | 35 (21.0%) | 19 (22.1%) | 15 (20.5%) | 0.966 |
| **Aspirin pre PPCI, n (%)** | 165 (98.8%) | 85 (98.8%) | 72 (98.6%) | 1.000 |
| **LAD culprit, n (%)** | 75 (44.9%) | 38 (44.2%) | 35 (47.9%) | 0.753 |
| **Multivessel disease >1 vessel disease, n (%)** | 49 (29.7%) | 25 (29.4%) | 22 (30.1%) | 1.000 |
| **TIMI flow pre PCI =0, n (%)** | 127 (76.0%) | 65 (75.6%) | 56 (76.7%) | 1.000 |
| **Thrombus score>2** | 141 (87.6%) | 76 (90.5%) | 61 (85.9%) | 0.528 |
| **TIMI flow post PCI =3, n (%)** | 145 (86.8%) | 77 (89.5%) | 62 (84.9%) | 0.527 |
| **Myocardial blush grade < 2, n (%)** | 20 (12.4%) | 8 (9.52%) | 11 (15.5%) | 0.377 |
| **Troponin peak level, ng/L, median [IQR]** | 49.4 [15.8; 90.6] | 41.3 [13.13; 50.0] | 54.5 [30.0-189] | **0.002** |
| **Creatinine level on admission, µmol/L, median [IQR]** | 75.0 [68.0; 87.5] | 77.0 [68.0; 86.8] | 74.0 [68.0-89.0] | 0.499 |
| **Peak monocyte count, x10<sup>9</sup>/L, median [IQR]** | 72.0 [62.0; 82.0] | 73.0 [63.8; 83.0] | 72.0 [62.0; 82.0] | 0.587 |
| **Peak neutrophils count, x10<sup>9</sup>/L, median [IQR]** | 9.41 [7.37; 11.7] | 8.86 [7.36; 11.6] | 9.73 [7.39; 11.8] | 0.686 |
| **ST-segment resolution**<br>**>70% on ECG, n (%)** | 125 (75.3%) | 61 (70.9%) | 56 (77.8%) | 0.426 |
| **Medications at discharge** |  |  |  |  |
| **Aspirin** | 162 (97.0%) | 83 (96.5%) | 71 (97.3%) | 1.000 |
| **Clopidogrel** | 63 (38.0%) | 25 (29.4%) | 31 (42.5%) | 0.123 |
| **Ticagrelor** | 102 (61.1%) | 60 (69.8%) | 41 (56.2%) | 0.107 |
| **Beta blockers** | 159 (95.2%) | 83 (96.5%) | 68 (93.2%) | 0.471 |
| **ACEI** | 157 (94.0%) | 80 (93.0%) | 70 (95.9%) | 0.509 |
| **ARB** | 9 (5.39%) | 5 (5.81%) | 3 (4.11%) | 0.727 |
| **Statin** | 165 (98.8%) | 85 (98.8%) | 72 (98.6%) | 1.000 |
| **Diuretics** | 14 (8.38%) | 9 (10.5%) | 4 (5.48%) | 0.394 |
| **Nitrates** | 142 (85.0%) | 74 (86.0%) | 62 (84.9%) | 1.000 |
| **Calcium Channel**<br>**blockers** | 11 (5.59%) | 6 (6.98%) | 5 (6.85%) | 1.000 |
| **Anti-coagulation** | 6 (3.59%) | 3 (3.49%) | 3 (4.11%) | 1.000 |
| **Medications at 6 months follow-up** |  |  |  |  |
| **Aspirin** | 143 (94.1%) | 75 (93.8%) | 61 (93.8%) | 1.000 |
| **Clopidogrel** | 61 (40.7%) | 24 (30.4%) | 30 (46.9%) | 0.064 |
| **Ticagrelor** | 86 (57.3%) | 54 (68.4%) | 32 (50.0%) | **0.040** |
| **Beta blockers** | 134 (88.7%) | 71 (89.9%) | 56 (86.2%) | 0.668 |
| **ACEI** | 125 (82.2%) | 64 (80.0%) | 56 (86.2%) | 0.451 |
| **ARB** | 21 (13.9%) | 11 (13.9%) | 8 (12.3%) | 0.970 |
| **Statin** | 146 (96.1%) | 77 (96.2%) | 62 (95.4%) | 1.000 |
| **Diuretics** | 12 (7.95%) | 7 (8.75%) | 4 (6.25%) | 0.755 |
| **Nitrates** | 117 (77.5%) | 59 (73.8%) | 55 (85.9%) | 0.113 |
| **Calcium Channel blockers** | 14 (9.27%) | 8 (10.1%) | 6 (9.23%) | 1.000 |
| **Anti-coagulation** | 4 (2.63%) | 2 (2.50%) | 2 (3.08%) | 1.000 |

At presentation, ID was prevalent, whatever definition was used. Indeed, 54.09% of patients had hypoferremia denoted as TSAT<20%, 60% of patients had hypoferremia denoted as serum iron<13μM, 45.5% of patients had hypoferritinemia, denoted as serum ferritin<100ug/L, and 69.1% of patients met the current ESC definition of ID. Of the 112 patients who met the ESC definition of ID at presentation, 82 (73.2%) had Tsat<20%. Unsurprisingly, relative to those with Tsat ≥20% at presentation, those with Tsat<20%, had lower ferritin (126 [71.2; 251]ug/L vs 97.0 [55.4; 166] ug/L, p=0.012), lower iron concentration (15.8 [11.4; 20.2] uM vs 9.06 [6.11; 12.1] uM, p<0.001), higher transferrin concentration (1.68 [1.44; 1.96] g/L vs 3.14 [2.24; 4.46] g/L, p<0.001), and were also more likely to meet the ESC definition of ID (37% vs 95.3%, p<0.001), or have serum iron <13uM (32.9% vs 80.2%, p<0.001). There were no differences between the two sub-groups in any demographics, risk factors, medications before STEMI or at discharge, or in clinical and angiographic characteristics, except for peak troponin levels, which were significantly lower in those with TSAT<20% than those with TSAT≥20% at presentation (41.3 [13.13; 50.0] ng/L vs 54.5 [30.0-189] ng/L, p=0.002). Ticagrelor use at 6 month follow-up was greater in those with TSAT<20% than those with TSAT≥20% at presentation (68.4% vs 50.0%, p=0.04).

### Hypoferremia at acute STEMI presentation is an independent predictor of lower risk of MACE

In the 163 patients with follow-up data, the median follow-up duration was 4.5 years [3.5; 6.0 years], and data were censored at the last day that information on MACE was available. During follow-up, 29 patients experienced a total of 34 MACE events; 25 patients experienced a single event, 3 patients experienced 2 events and 1 patient experienced three events. The 34 MACE events comprised 4 cardiac deaths, 17 recurrent MI and 13 new diagnoses of HF.

First, we examined differences in probability of MACE-free survival between those with or without ID at presentation. Kaplan-Meier survival plots are shown in figure 2. The presence of ID at presentation was associated with a higher probability of MACE-free survival, whether ID was defined as TSAT<20% (p=0.027), as serum iron <13uM (p=0.017) or according to ESC definition (p=0.0092). The presence of hypoferritinemia (as ferritin<100ug/L) at presentation was not associated with any difference in probability of MACE-free survival (supplemental figure 1). Markers of ID sampled either immediately after the PCI procedure or 6 hours post PCI were still associated with a higher probability of MACE-free survival. In contrast, the association between ID and MACE-free survival was not observed when iron markers were sampled at timepoints later than 6 hours after PCI.

**Figure 2.**
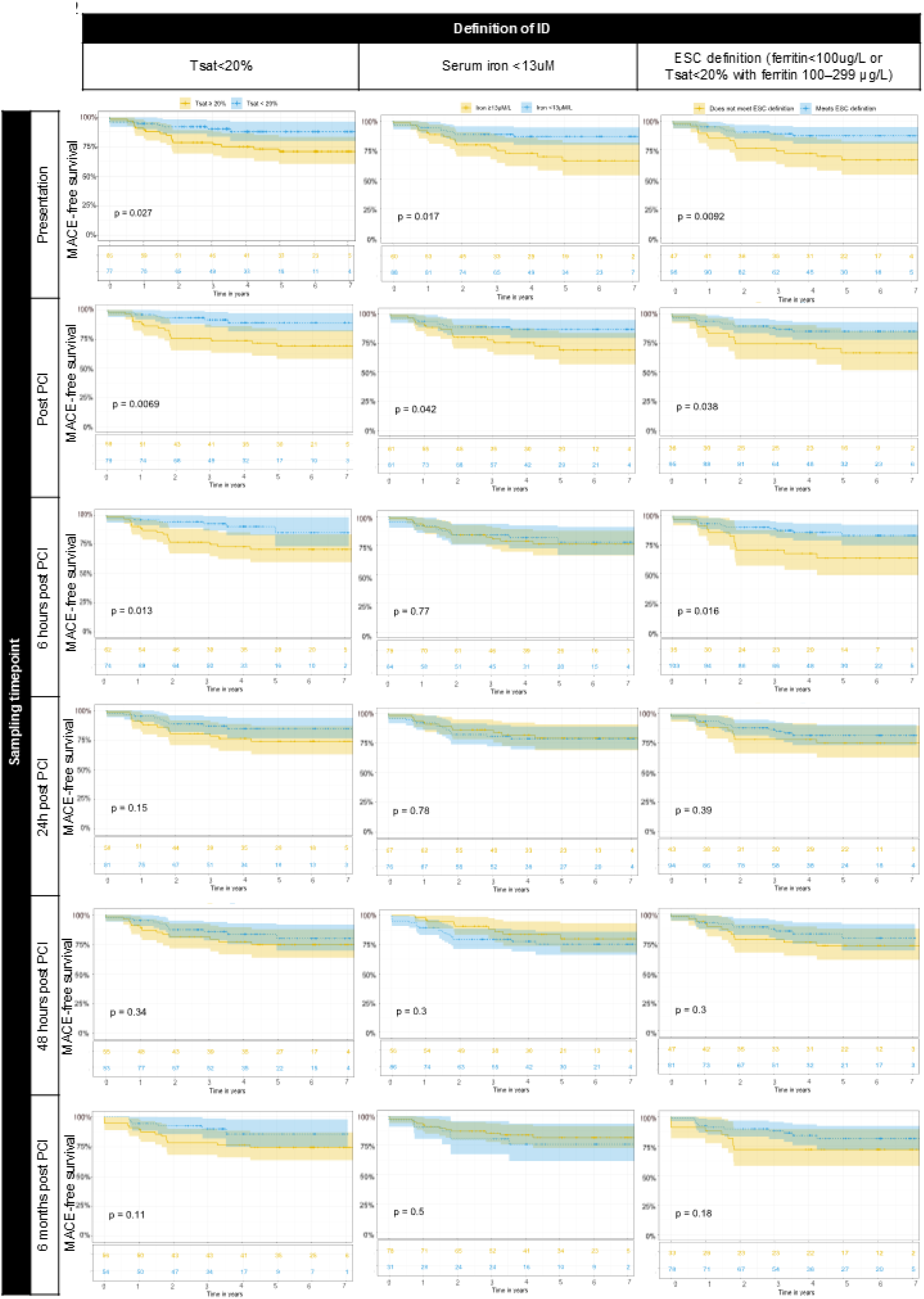
Hypoferremia at acute STEMI presentation is associated with better MACE-free survival. Kaplan Meier survival plots showing percentage MACE-free survival. For each plot, patients were stratified according to whether they meet, or not, a particular definition of iron deficiency, Tsat<20% (left), serum iron <13uM (centre), or ESC definition of iron deficiency (right). Patients were re-stratified in this manner using Tsat, iron and ferritin values sampled either at presentation (top row), immediately post PCI (second row), 6 hours post PCI (third row), 24 hours post PCI (fourth row), 48 hours post PCI (fifth row) or at 6 month follow-up (bottom row). P values shown were calculated using long rank test.

We next performed multivariate Cox regression to estimate the effect of ID at presentation on long-term risk of MACE, using alternative definitions of ID. In each case, we corrected for age, gender, BMI, ACEI/ARB, Beta blocker or Statin use pre STEMI, smoker status, hypertension, diabetes, hypercholesterolemia, family history of CAD, COPD or asthma, previous PCI and previous MI. Figure 3 shows a summary forest plot of adjusted hazard ratios, estimated via separate cox regression analyses, for each sampling timepoint and each definition of ID. The risk of long-term MACE was lower for those with ID at presentation, when ID was defined as Tsat<20% [HR=0.38 (CI 0.16-0.90), p=0.028 relative to Tsat≥20%], according to ESC definition of ID (HR=0.36, CI [0.16-0.82], p=0.015) relative to not meeting ESC definition), or as serum iron <13uM (HR=0.41, CI [0.19-0.89], p=0.023) relative to iron≥13uM). The risk of long-term MACE was not different between strata when ID was defined as ferritin<100ug/L (supplemental figure 2). When we repeated this analysis using iron markers sampled at later timepoints, Tsat<20% immediately after or 6 hours after PCI, or having iron<13µM immediately after PCI still predicted lower risk of MACE. The complete forest plots comprising other co-variates are shown in supplemental figures 3, 4 and 5.

**Figure 3.**
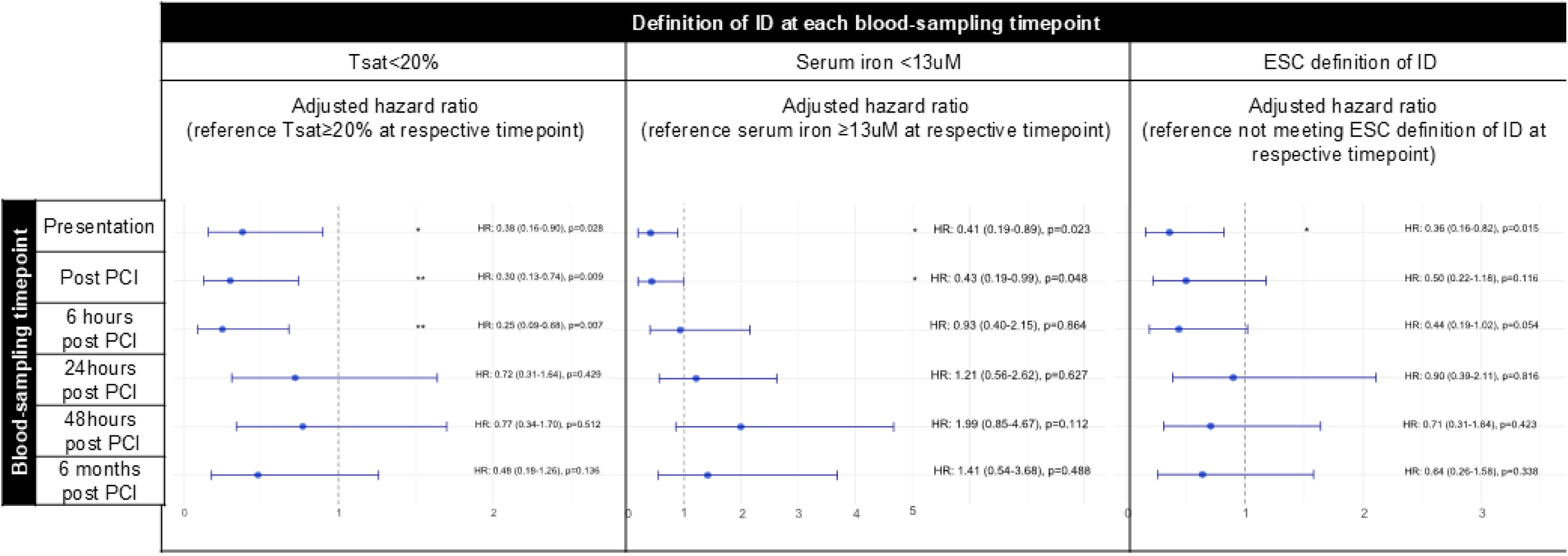
Hypoferremia at acute STEMI presentation is an independent predictor of lower risk of MACE. Summary forest plot of adjusted hazard ratios for MACE associated with each definition of ID, at each sampling timepoint. Cox regression analysis for risk of MACE was based on the most parsimonious model. Modelling was performed separately for each sampling timepoint. Complete forest plots with other co-variates are shown in supplemental figures 3 and 4

These findings demonstrate that hypoferremia, but not hypoferritinemia, at acute STEMI presentation predicts lower long-term risk of MACE. They also show that this predictive effect is lost when iron markers are sampled at later timepoints, likely because these markers change over time secondary to STEMI and or clinical intervention, consequently becoming less reflective of a patient’s baseline status (supplemental figure 6).

### Hypoferremia at presentation predicts attenuated acute myocardial injury and improved early LV remodelling

Long-term outcomes after STEMI reflect the culmination of processes that evolve in the hours, days, weeks and months post STEMI; from the acute myocardial injury, and early LV remodelling to subsequent cardiac salvage, to late LV remodelling. We considered the possibility that baseline hypoferremia predicted long-term outcomes by favourably impacting some of these processes. The observation that those with TSAT<20% at presentation have significantly lower peak troponin levels than those with baseline TSAT≥20% (41.3 [13.13; 50.0] ng/L vs 54.5 [30.0-189] ng/L, p=0.002), point towards the hypothesis that a patient’s iron status could influence the extent of myocardial injury. To test this hypothesis, we used regression modelling to estimate the effect of Tsat on acute CMR parameters and other relevant plasma markers of myocardial injury and early remodelling measured before hospital discharge. Relative effect sizes and the direction of effects are represented as a heatmap (Figure 4A). For markers that were significantly affected by Tsat, slopes generated by prediction models are shown in separate plots (Figure 4B). All effect size estimates and p values are reported in table 2.

**Figure 4.**
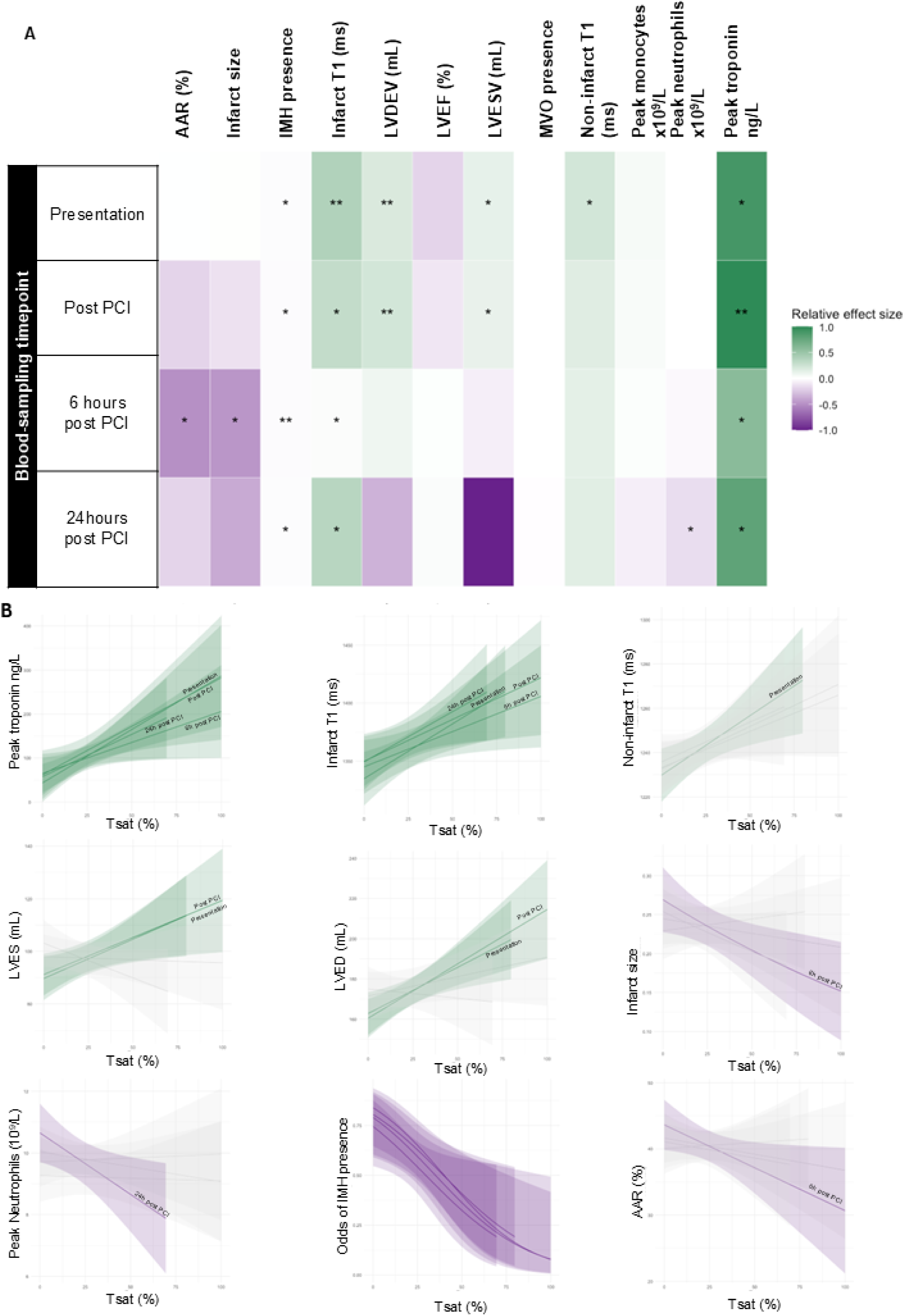
Hypoferremia at presentation predicts attenuated acute myocardial injury and improved early LV remodelling. A. Heatmap representing relative adjusted effect sizes of Tsat (per unit, 1 %) on CMR variables and other serum markers collected before hospital discharge. Positive effects are shown in green, and negative effects shown in purple, with the depth of the colour denoting the relative size of the effect. Regression models were run separately for each sampling timepoint*p<0.5, **p<0.01. B. Prediction plots for adjusted effect sizes of Tsat on acute CMR variables and other markers of myocardial injury collected before hospital discharge. Plots only shown for CMR and serum variables that were significantly associated with Tsat. Sampling timepoints that did not have significant effects are shaded in grey.

**Table 2.**
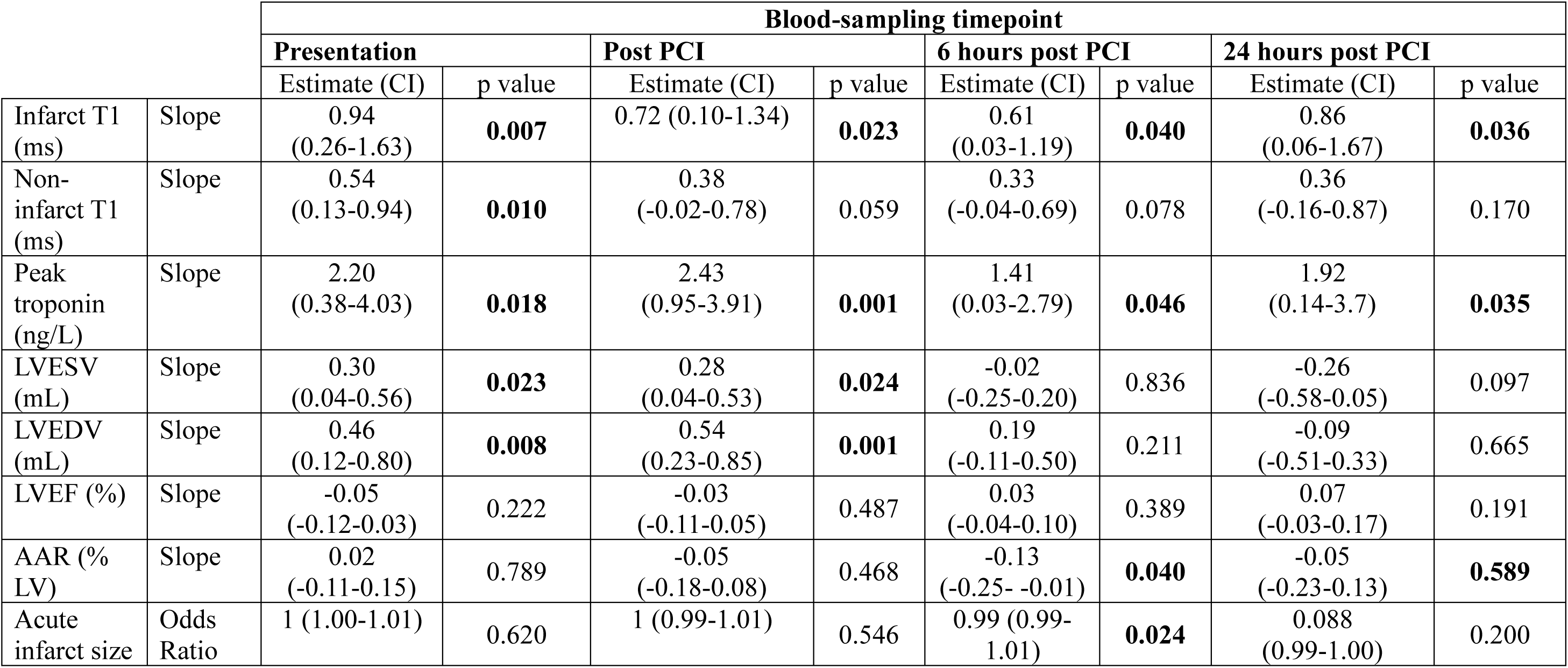
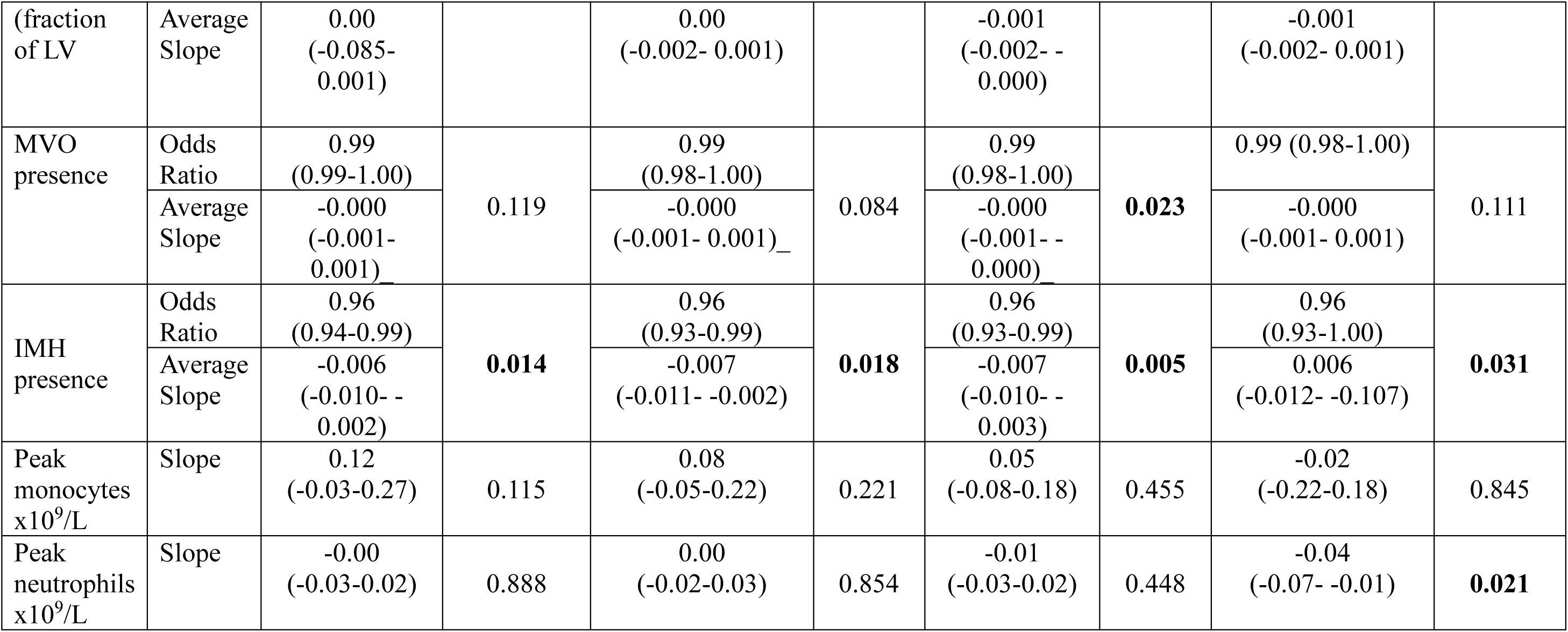
Regression Modelling of the effect of Tsat on acute CMR parameters and blood markers collected before discharge.

After controlling for all confounders, Tsat at presentation predicted higher acute infarct T1, non-infarct T1, peak Troponin; all markers of acute myocardial injury. Indeed, every one unit (1%) increase in Tsat at presentation predicted 0.94ms higher infarct T1 (CI 0.26-1.63, p=0.007), 0.5ms higher non-infarct T1 (CI [0.13-0.94], p=0.01), 2.2ng/L higher peak troponin (CI [0.3-4.03], p=0.018). Tsat at presentation also predicted higher LVEDV and LVESV; markers of adverse early LV remodelling. Indeed, every one unit (1%) increase in Tsat at presentation predicted 0.46mL higher LVEDV (CI [0.12-0.8], p=0.008) and 0.3mL higher LVESV (CI [0.04-0.56], p=0.023). When this analysis was repeated for Tsat sampled at later timepoints, the predictive effect on non-infarct T1 was lost, on infarct T1 and peak Troponin was sustained up to 24 hours post PCI, and on LVES and LVED was sustained at immediately post PCI only (Figure 4). Conversely, every one unit (1%) increase in Tsat at presentation predicted 4% (CI [1-6], p=0.014) lower odds of intramyocardial haemorrhage (IMH) and this predicted effect was sustained for Tsat sampled up to 24 hours post PCI. Furthermore, every one unit increase in Tsat at 6 hours post PCI (but not at presentation) predicted 0.1% smaller acute infarct size (CI [−0.2% – −0.0%], p=0.024), and 0.13% smaller acute area at risk (CI [−0.25% – −0.01], p=0.04). Every one unit increase in Tsat at 24h post PCI predicted 0.04 x10^9^/L lower levels of peak circulating neutrophil counts (CI [−0.0 – −0.01], p=0.21).

These data indicate that hypoferremia at presentation predicts multiple aspects of the early response to STEMI, on the one hand predicting attenuated acute myocardial injury to the infarct and remote myocardium, and improved early LV remodelling, and on the other hand higher odds of IMH.

### Hypoferremia at presentation is associated with lower myocardial salvage at 6 months

We next examined the relationship between hypoferremia and later processes that mediate myocardial repair and late LV remodelling after STEMI. To that effect, we used regression modelling to estimate the effect of Tsat on CMR parameters and NT-pro-BNP levels measured at 6 months. Baseline characteristics of patients who returned for the 6-month scan are shown in table 3. Relative effect sizes and the direction of effects are represented as a heatmap (Figure 5A). For markers that were significantly affected by Tsat, slopes generated by prediction models are shown in separate plots (Figure 5B). All effect size estimates are reported in table 4.

**Table 3.** Baseline characteristics of patients who returned for 6 month scan.

|  | **All Cohort**<br>**(N=167)** | **Did not return**<br>**for 6-month**<br>**scan**<br>**(N=46)** | **Returned for 6-**<br>**month scan**<br>**(N=121)** | **P value** |
| --- | --- | --- | --- | --- |
| **Iron status at admission** |  |  |  |  |
| **Ferritin (µg/L), median [IQR]** | 110 [64.8; 193] | 114 [61.6; 193] | 107 [68.7; 194] | 0.656 |
| **Ferritin<100µg/L , n (%)** | 75 (45.5%) | 19 (42.2%) | 56 (46.7%) | 0.738 |
| **Plasma iron (µM), median [IQR]** | 11.4 [7.22; 16.7] | 11.2 [5.86; 16.9] | 11.4 [7.33; 15.7] | 0.430 |
| **Plasma iron <13Mm, n (%)** | 99 (60.0%) | 25 (55.6%) | 74 (61.7%) | 0.592 |
| **Plasma transferrin (g/L) median [IQR]** | 2.08 [1.64; 3.32] | 2.04 [1.72; 2.52] | 2.10 [1.64; 3.33] | 0.802 |
| **ESC definition of ID (Ferritin<100µg/L or 100–299 µg/L with TSAT < 20%), n (%)** | 112 (69.1%) | 29 (67.4%) | 83 (69.7%) | 0.930 |
| **Demographics and risk factors** |  |  |  |  |
| **Gender, male, n (%)** | 141 (84.9%) | 40 (88.9%) | 101 (83.5%) | 0.533 |
| **Age, median [IQR]** | 61.0 [54.0; 68.0] | 59 [52.0; 67.0] | 62.0 [54.0; 68.0] | 0.271 |
| **BMI, median [IQR]** | 27.2 [24.6; 29.9] | 27 [24.8; 30.7] | 27.3 [24.5; 29.5] | 0.556 |
| **Smoking history, n (%)** | 113 (68.1%) | 34 (75.6%) | 79 (65.3%) | 0.283 |
| **Hypertension, n (%)** | 74 (44.3%) | 21 (45.7%) | 53 (43.8%) | 0.968 |
| **Diabetes, n (%)** | 35 (21.0%) | 17 (37.0%) | 18 (14.9%) | **0.004** |
| **Hypercholesterolemia, n (%)** | 59 (35.3%) | 18 (39.1%) | 41 (33.9%) | 0.651 |
| **Family history of CAD, n (%)** | 82 (49.4%) | 22 (48.9%) | 60 (49.6%) | 1.000 |
| **Previous MI, n (%)** | 9 (5.39%) | 4 (8.70%) | 5 (4.13%) | 0.262 |
| **Previous PCI, n (%)** | 6 (3.66%) | 3 (6.82%) | 3 (2.50%) | 0.084 |
| **COPD or Asthma, n (%)** | 18 (10.8%) | 6 (13.0%) | 12 (9.9%) | 0.581 |
| **Medications before STEMI** |  |  |  |  |
| **ACEI or ARB, n (%)** | 32 (19.2%) | 12 (26.1%) | 20 (16.5%) | 0.237 |
| **Beta blocker, n (%)** | 17 (10.2%) | 8 (17.4%) | 9 (7.44%) | 0.083 |
| **Statin, n (%)** | 34 (20.4%) | 11 (23.9%) | 23 (19.0%) | 0.625 |
| **Clinical and angiographic characteristics** |  |  |  |  |
| **BP systolic on admission mmHg, median [IQR]** | 128 [111; 150] | 126 [114; 150] | 128 [111; 149] | 0.969 |
| **HR on admission beats/min, median [IQR]** | 80 [65; 90] | 79 [64.2; 86.5] | 80.0 [68.0; 90.0] | 0.439 |
| **Pain to PCI time (min), mean [IQR]** | 189 [125; 298] | 220 [126; 388] | 175 [124; 272] | 0.238 |
| **Late presenter (>6h), n (%)** | 35 (21.0%) | 14 (30.4%) | 21 (17.4%) | 0.100 |
| **Aspirin pre PPCI, n (%)** | 165 (98.8%) | 45 (97.8%) | 120 (99.2%) | 0.476 |
| **LAD culprit, n (%)** | 75 (44.9%) | 17 (37%) | 58 (47.9%) | 0.271 |
| **Multivessel disease >1 vessel disease, n (%)** | 49 (29.7%) | 13 (28.9%) | 36 (30.0) | 1.000 |
| **TIMI flow pre PCI =0, n (%)** | 127 (76.0%) | 36 (78.3%) | 91 (75.2%) | 0.833 |
| **Thrombus score>2** | 141 (87.6%) | 37 (88.1%) | 104 (87.4%) | 1.000 |
| **TIMI flow post PCI =3, n (%)** | 145 (86.8%) | 40 (87.0%) | 105 (86.8%) | 1.000 |
| **Myocardial blush grade < 2, n (%)** | 20 (12.4%) | 3 (7.14%) | 17 (14.3%) | 0.350 |
| **Troponin peak level, ng/L, median [IQR]** | 49.4 [15.8; 90.6] | 50.0 [13.3;70.1] | 47.9 [17.8; 97.9] | 0.910 |
| **Creatinine level on admission, µmol/L, median [IQR]** | 75.0 [68.0; 87.5] | 79 [69.0; 94.0] | 75 [68.0; 86.0] | 0.201 |
| **Peak monocyte count, x10<sup>9</sup>/L, median [IQR]** | 72.0 [62.0; 82.0] | 73.0 [68.0; 84.0] | 69.5 [61.2; 81.8] | 0.131 |
| **Peak neutrophils count, x10<sup>9</sup>/L, median [IQR]** | 9.41 [7.37; 11.7] | 8.23 [7.10; 11.6] | 9.74 [7.33; 11.8] | 0.269 |
| **ST-segment resolution >70% on ECG, n (%)** | 125 (75.3%) | 35 (77.8%) | 90 (74.4%) | 0.804 |
| **Medications at discharge** |  |  |  |  |
| **Aspirin** | 162 (97.0%) | 44 (95.7%) | 118 (97.5%) | 0.616 |
| **Clopidogrel** | 63 (38.0%) | 18 (40.0%) | 45 (37.2%) | 0.879 |
| **Ticagrelor** | 102 (61.1%) | 27 (58.7%) | 75 (62.0%) | 0.832 |
| **Beta blockers** | 159 (95.2%) | 44 (95.7%) | 115 (95.0%) | 1.000 |
| **ACEI** | 157 (94.0%) | 40 (87.0%) | 117 (96.7%) | **0.027** |
| **ARB** | 9 (5.39%) | 5 (10.9%) | 4 (3.31%) | 0.117 |
| **Statin** | 165 (98.8%) | 45 (97.8%) | 120 (99.2%) | 0.476 |
| **Diuretics** | 14 (8.38%) | 4 (8.70%) | 10 (8.26%) | 1.000 |
| **Nitrates** | 142 (85.0%) | 36 (78.3%) | 106 (87.6%) | 0.204 |
| **Calcium Channel blocker** | 11 (5.59%) | 2 (4.35%) | 9 (7.44%) | 0.729 |
| **Anti-coagulation** | 6 (3.59%) | 1 (2.17%) | 5 (4.13%) | 1.000 |
| **Medications at 6 months follow-up** |  |  |  |  |
| **Aspirin** | 143 (94.1%) | 31 (93.9%) | 112 (94.1%) | 1.000 |
| **Clopidogrel** | 61 (40.7%) | 16 (51.6%) | 45 (37.8%) | 0.235 |
| **Ticagrelor** | 86 (57.3%) | 13 (41.9%) | 73 (61.3%) | 0.081 |
| **Beta blockers** | 134 (88.7%) | 29 (90.6%) | 105 (88.2%) | 1.000 |
| **ACEI** | 125 (82.2%) | 27 (81.8%) | 98 (82.4%) | 1.000 |
| **ARB** | 21 (13.9%) | 4 (12.5%) | 17 (14.3%) | 1.000 |
| **Statin** | 146 (96.1%) | 31 (93.9%) | 115 (96.6%) | 0.611 |
| **Diuretics** | 12 (7.95%) | 3 (9.38%) | 9 (7.56%) | 0.718 |
| **Nitrates** | 117 (77.5%) | 17 (53.1%) | 100 (84.0%) | **0.001** |
| **Calcium**<br>**Channel blockers** | 14 (9.27%) | 1 (3.12%) | 13 (10.9%) | 0.303 |
| **Anti-coagulation** | 4 (2.63%) | 1 (3.03%) | 3 (2.52%) | 1.000 |

**Figure 5.**
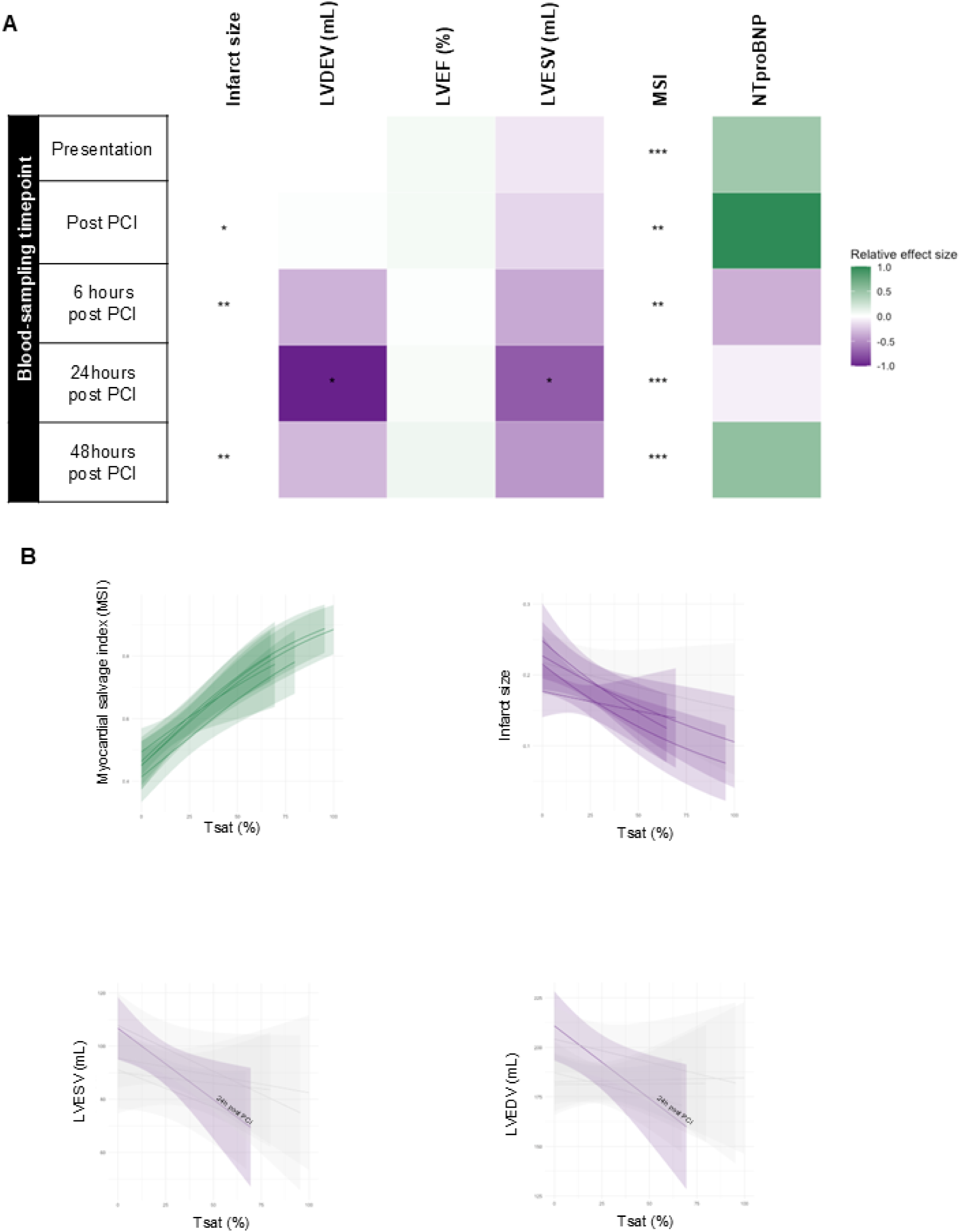
Hypoferremia at presentation is associated with lower myocardial salvage at 6 months. A. Heatmap representing relative adjusted effect sizes of Tsat (per unit, 1 %) on CMR variables and NTproBNP at 6 months. Positive effects are shown in green, and negative effects shown in purple, with the depth of the colour denoting the size of the effect. Regression models were run separately for each sampling timepoint. *p<0.5, **p<0.01, ***p<0.001 B. Prediction plots for adjusted effect sizes of Tsat on CMR variables and other serum markers of cardiac remodelling collected at 6 months post discharge. Plots only shown for CMR and serum variables that were significantly associated with Tsat. Sampling timepoints that did not have significant effects are shaded in grey.

**Table 4.** Regression Modelling of the effect of Tsat on 6 month CMR and NT-proBNP.

|  |  | Timepoint of Tsat sampling |  |  |  |  |  |  |  |  |  |
| --- | --- | --- | --- | --- | --- | --- | --- | --- | --- | --- | --- |
|  |  | Presentation |  | Post PCI |  | 6 hours post PCI |  | 24 hours post PCI |  | 48 hours post PCI |  |
|  |  | Estimate (CI) | p value | Estimate (CI) | p value | Estimate (CI) | p value | Estimate (CI) | p value | Estimate (CI) | p value |
| LVESV (mL) | Slope | -0.08<br>(-0.48-0.32) | 0.683 | -0.13<br>(-0.50-0.25) | 0.494 | -0.28<br>(-0.66-0.09) | 0.138 | -0.54<br>(-0.97- -0.10) | **0.017** | -0.34<br>(-0.72-0.03) | 0.069 |
| LVEDV (mL) | Slope | 0.01<br>(-0.51-0.52) | 0.975 | 0.02<br>(-0.47-0.51) | 0.933 | -0.25 (-0.76-0.27) | 0.346 | - 0.73 (-1.31- -0.15) | **0.014** | -0.23 (-0.75-0.28) | 0.370 |
| LVEF (%) | Slope | 0.07<br>(-0.02-0.16) | 0.122 | 0.07<br>(-0.02-0.17) | 0.128 | 0.02<br>(-0.09-0.13) | 0.739 | 0.05<br>(-0.06-0.17) | 0.364 | 0.10<br>(-0.01-0.20) | 0.065 |
| Infarct size (fraction of LV) | Odds Ratio | 1.00<br>(0.99-1.00) | 0.295 | 0.99<br>(0.98-1.00) | **0.038** | 0.99<br>(0.98-1.00) | **0.008** | 1.00<br>(0.98-1.01) | 0.466 | 0.99<br>(0.98-1.00) | **0.009** |
|  | Average Slope | -0.001<br>(-0.002-0.001) |  | -0.001<br>(-0.002- -0.001) |  | -0.002<br>(-0.003- -0.001) |  | -0.001<br>(-0.002-0.001) |  | -0.002<br>(-0.003- -0.001) |  |
| MSI (fraction of LV mass) | Odds Ratio | 1.02<br>(1.01-1.03) | <**0.001** | 1.02<br>(1.01-1.03) | <**0.001** | 1.02<br>(1.01-1.04) | <**0.001** | 1.02 (1.00-1.03_ | **0.012** | 1.02 (1.01-1.03) | <**0.001** |
|  | Average Slope | 0.005<br>(0.002 – 0.007) |  | 0.005<br>(0.002 – 0.007) |  | 0.005<br>(0.002 – 0.008) |  | 0.005<br>(0.001 – 0.008) |  | 0.005<br>(0.002 – 0.007) |  |
| NTproBNP ng/L |  | 0.57<br>(-4.59-5.73) | 0.826 | 1.20<br>(-3.45-5.84) | 0.609 | -0.26<br>(-5.13-4.62) | 0.916 | -0.05<br>(-5.95-5.85) | 0.987 | 0.66<br>(-4.57-6.07) | 0.809 |

After correcting for all confounders, Tsat at presentation predicted higher myocardial salvage index (MSI), with every one unit (1%) increase in Tsat predicting 0.5% (CI [0.2% – 0.7%], p<0.001) higher MSI. This effect was sustained for Tsat sampled immediately after and up to 48 hours post PCI. Moving away from baseline, Tsat sampled immediately post PCI predicted lower infract size, with every one unit (1%) increase in post PCI Tsat predicting 0.1% (CI [−0.2% – −0.1%], p=0.038) lower infarct size. Tsat at 24 hours post PCI predicted lower LVEDV and LVES, with every one unit increase (1%) in Tsat at 24 hours predicting 0.76mL (CI [−1.31 – −0.15], p=0.014) lower LDVED and 0.54mL (CI [−0.97 – −0.01], p=0.017) lower LVES. These data demonstrate that hypoferremia at presentation is associated with impaired myocardial repair.

### MI in mice triggers an acute reactive iron in the myocardium, early LV remodelling and progression to HF, all of which are averted by dietary iron restriction

To gain greater mechanistic insights into the clinical findings, we utilised a well-established standard mouse model of MI, induced surgically by ligation of the left ascending artery (LAD) (25). SHAM-operated controls underwent an identical surgery except for the ligation step. Mice were placed on a standard diet or an iron-restricted diet 6 to7 weeks prior to surgery and maintained on those respective diets throughout.

As a first step, we sought to ascertain whether this mouse model of MI recapitulated the favourable effect of low iron levels on risk of MACE observed in STEMI patients. To that effect, we assessed the incidence of HF, defined here as LVEF≤45%, at 10 weeks post surgery. In the MI group, 12/20 of the mice receiving a standard iron diet developed HF, whereas only 1/8 mice receiving an iron-restricted diet developed HF (p=0.0377). None of the mice in the SHAM groups developed HF at 10 weeks (Figure 6A). In mice on standard iron diet, mean LVEF at 10 weeks was lower in the MI group than in respective SHAM controls (42.2±3.0% vs 69.7±6.6%, p<0.0001). In mice on an iron-restricted diet, there was no significant difference in LVEF between the MI group and SHAM controls (64.4±1.6% vs 72.7±1.8%, p=0.534). However, mean LVEF was significantly lower in the MI group on the standard iron diet than in the MI group on the iron-restricted diet (44.2±3.0% vs 64.4±1.6%, p=0.0011) (Figure 6B). In mice on standard iron diet, MI resulted in significant LV remodelling compared to SHAM controls, manifesting in higher LVEDV (81.1±9.6ul vs 47.3±4.1ul, p=0.009) and higher LVESV (46.07±7.7ul vs 14.4±1.5ul, p=0.0118). In mice on an iron-restricted diet, MI did not trigger significant LV remodelling compared to SHAM controls in terms of LVEDV (51.6±7.6ul vs 49.8±4.0ul, p=0.922) and LVES (20.18±5.1 vs 13.78±1.7ul, p=0.956) (Figure 6C, D).

**Figure 6.**
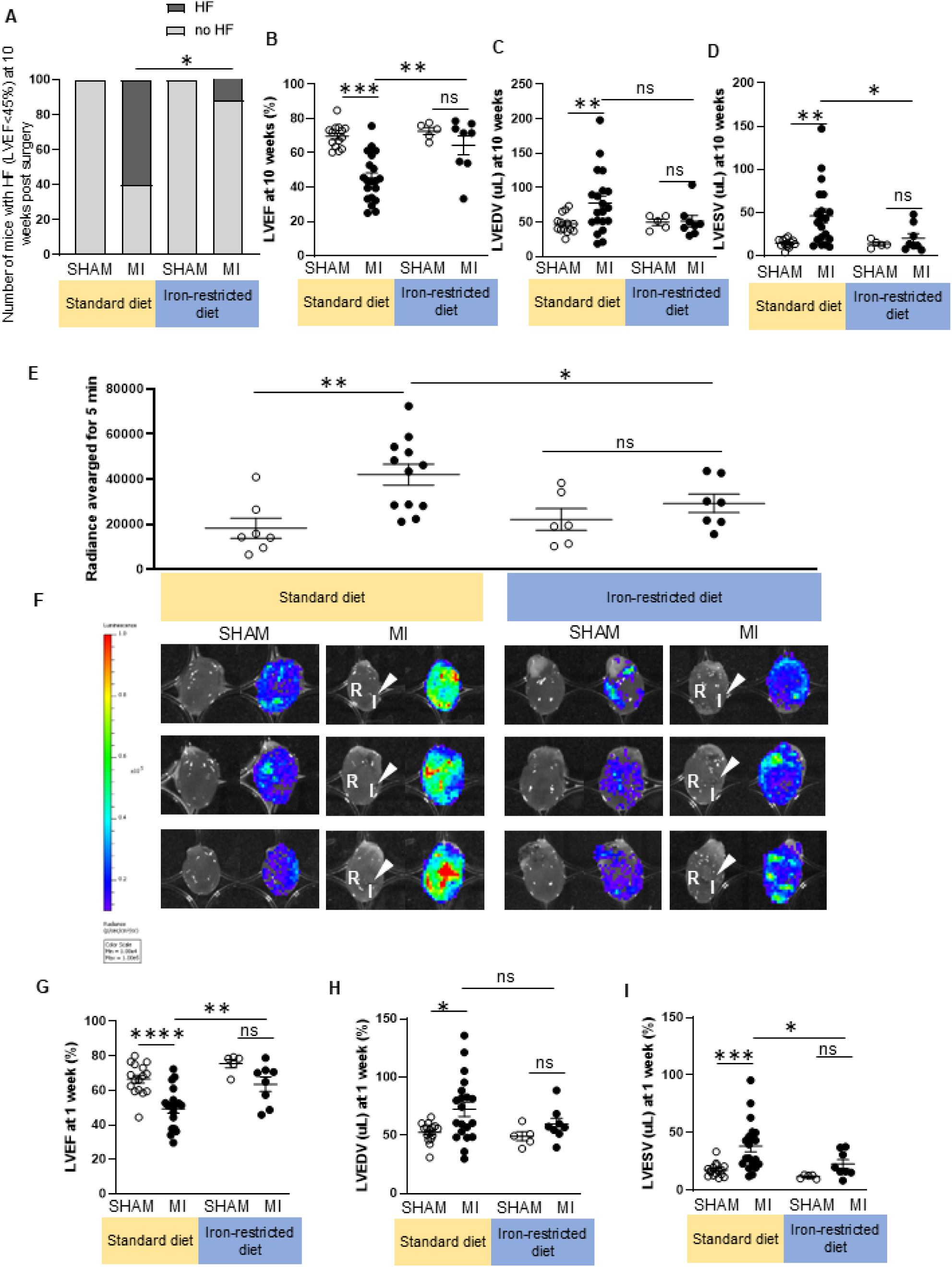
MI in mice triggers an acute reactive iron in the myocardium, early LV remodelling and progression to HF, all of which are averted by iron-restricted diet. A. Incidence of HF (LVEF≤45%) at 10 weeks, in mice that underwent MI or SHAM controls, maintained on a standard iron or iron-restricted diet throughout. B. LVEF at 10 weeks, in mice that underwent MI or SHAM controls, maintained on a standard iron or iron-restricted diet throughout. C. LVEDV at 10 weeks, in mice that underwent MI or SHAM controls, maintained on a standard iron or iron-restricted diet throughout. D. LVESV at 10 weeks, in mice that underwent MI or SHAM controls, maintained on a standard iron or iron-restricted diet throughout. E. Luminescence-based quantitation of intracellular reactive iron in the hearts of mice, 6 hours after MI and in SHAM controls. F. Representative images of mouse hearts, without luminescence (left) and with luminescence signal super-imposed (right). Arrows denote the site of LAD ligation. “I” denotes the infarcted area, discerned visually as discoloration downstream of the site of LAD ligation. “R” denotes the area remote to the site of the infarct. G. LVEF at 1 week, in mice that underwent MI or SHAM controls, maintained on a standard iron or iron-restricted diet throughout. H. LVEDV at 1 week, in mice that underwent MI or SHAM controls, maintained on a standard iron or iron-restricted diet throughout. I. LVESV at 1 week, in mice that underwent MI or SHAM controls, maintained on a standard iron or iron-restricted diet throughout. ns= not significant, *p<0.5, **p<0.01, ***p<0.001

Having established that the preclinical model recapitulates long-term clinical outcomes, we set out to evaluate the mechanisms underlying the effects of iron status on myocardial injury. Because myocardial injury is driven by oxidative stress, and intracellular reactive iron is a potent catalyst for oxidative stress, we assessed the impact of MI and of iron status on levels of intracellular reactive iron within the myocardium, using luminescence reporter luciferase and an iron-caged luciferin substrate. Quantitation of reactive-iron luminescence is shown in figure 6E. Representative luminescence images of mouse hearts are shown in figure 6F. In each case, an accompanying non-luminescence photograph is included, with the site of LAD ligation denoted with a white arrow, and the infarct and remote myocardium denoted by the letters “I” and “R” respectively. In mice on standard iron diet, MI triggered a rapid rise in intracellular reactive iron compared to SHAM controls, and this rise occurred both in the infarct and remote myocardium (Figure 6F). In mice on an iron-restricted diet, MI did not trigger this reactive iron storm (Figure 6E, F). These data mirror the clinical finding that Tsat at presentation was positively associated with both infarct T1 and non-infarct T1 at 2 days. We also evaluated early LV remodelling at 1 week post MI. In mice on standard iron diet, MI triggered an increase in LVEDV (72.2±6.0ul vs 52.4±3.3ul, p=0.0126), and LVES (38.3±4.8ul vs 17.5±5.5 ul, p=0.0005) and decline in LVEF (49.4±2.55% vs 66.5±8.77%, p<0.0001) compared to SHAM controls on the respective diet. In mice maintained on an iron-restricted diet, there were no significant differences between MI mice and SHAM controls. Comparing the extent of early LV remodelling in MI mice at 1 week, those on a standard iron diet had significantly higher LVES (38.3±4.7ul vs 22.64±3.6ul, p=0.0488), and significantly lower LVEF (49.4±2.55% vs 63.52±3.89%, p=0.0069) those on an iron-restricted diet, though there were no significant differences between the dietary groups in terms of LVEDV at this stage.

These preclinical data show MI triggers a myocardial reactive iron storm and that hypoferremia prevents this storm. These findings provide a mechanistic explanation for the clinical finding that hypoferremia at presentation predicts lower acute myocardial injury in patients with acute STEMI.

## Discussion

The central discovery of the present study is that hypoferremia at presentation with acute STEMI reduces long-term risk of MACE. This discovery is based on the longest period of follow-up for outcomes by iron status reported to date. The present study reconciles the seemingly conflicting results of previous studies by resolving key unknowns. First, it applied different definitions of ID, revealing that hypoferremia, but not hypoferritinemia predicts long-term outcomes. Second, it compared different sampling timepoints, illustrating how rapid and transient changes secondary to STEMI affect the results of analysis. Third, it evaluated multiple parameters of the cardiac response to STEMI, revealing that hypoferremia affects some processes favourably, and others adversely. Nonetheless, the sum of these effects is ultimately better long-term clinical outcomes (graphical abstract).

The finding that hypoferremia but not hypoferritinemia predicts risk of MACE indicates that it is iron in the serum or in the myocardium itself, at the point at which STEMI occurs, that ultimately influences outcomes. The roles of iron as a catalyst for oxidative damage, as a trigger for cardiomyocyte death, and as a fuel for IRI in the context of MI are already well established (10, 13–15). What this study reveals, for the first time, is that those roles do ultimately translate into adverse long-term clinical outcomes.

The preclinical model of MI not only corroborated these clinical findings, but also provided mechanistic insight into the impact of iron status on acute myocardial injury. It revealed that MI rapidly triggers a reactive iron storm in the myocardium. One potential pathway through which MI could trigger this reactive iron storm is ferritinophagy, when local ferritin stores inside cardiomyocytes are degraded, unlocking reactive iron within them. This process was previously demonstrated to account for a rise in myocardial reactive iron and oxidative stress and to contribute to the development of HF in a mouse model of pressure overload (26).

Additionally, inhibition of ferritinophagy in rats was shown to reduce oxidative stress and to ameliorate IRI after MI (27). Previous work published by this team demonstrated that provision of an iron-restricted diet reduced local myocardial ferritin stores in mice (21). Thus, mice on an iron-restricted diet have less fuel for ferritinophagy, potentially explaining why MI did not trigger a myocardial reactive iron storm in this setting. Whether hypoferremia in patients is also accompanied by a reduction in myocardial ferritin remains unknown. Further research is needed to unravel how different iron species are perturbed in the myocardium after MI.

Acute clinical CMR data revealed that hypoferremia at presentation is associated with lower T1, both in the infarct and remote myocardium, and with lower LVES and LVED. Mirroring this finding, provision of an iron-restricted diet in mice averted the reactive iron storm after MI, both in the infarcted and remote myocardium, and also prevented the early rise in LVES and LVED at 1 week. Elevated acute myocardial T1 mapping after STEMI is a known predictor of higher long-term risk MACE (22, 28). Equally, early LV remodelling, denoted by rise in LVEDV at 6 days post STEMI was previously found to predict higher risk of MACE over a follow-up period of ~ 6 years (29). Taken together, these lines of evidence converge on the notion that hypoeferremia improves long-term outcomes by attenuating the initial myocardial injury and mitigating the early LV remodelling. Mechanistic examination of 6-month CMR data, on the other hand, revealed that hypoferremia at presentation predicted lower MSI at 6 month follow-up. One limitation of MSI is that it is a function of both the extent of initial myocardial injury and of subsequent repair, with iron potentially influencing both processes. Nonetheless, the observation of ID is associated with lower MSI echoes findings by other (4) and aligns with a known role for iron in cardiac repair processes that take place in the days and weeks post MI (16–18).

In terms of future translation, this research highlights how iron could be manipulated to improve long-term outcomes post STEMI. For instance, a stepwise approach could be developed involving two interventions, first chelation of reactive before PCI, to minimise myocardial injury and early LV remodelling, followed, days later, by iron supplementation to promote myocardial salvage. These interventions have been shown, separately, to hold promise. In canines, administration of the iron chelator deferoxamine (DFO) in the immediate aftermath of coronary occlusion improves myocardial energetics and decreases infarct size (30,31). In patients, DFO administration before PCI was found to reduce oxidative stress and IRI (32,33). Equally, IV iron oxide (ferumoxytol) administration at 4 days post STEMI was found to be associated with greater MSI and better LV function at 3-month follow-up (34). In an anaemic mouse model of MI, deferring intravenous iron supplementation from 1 hour to 24 hours post MI, reversed LV remodelling, and improved cardiac contractility (35). Both interventions are subject to ongoing clinical trials. The MIRON-DFP trial will deliver deferiprone acutely to remove iron from the myocardium (36). The INFERRCT trial, will deliver IV iron between 3 days and 4 weeks post MI. Based on the present data, this timing is optimal as it is likely to favour beneficial effects by improving cardiac repair (37).

### Study strengths and limitations

The study strengths lie in the extended length of follow-up for MACE, the evaluation of different definitions of ID and at multiple sampling timepoints, in mechanistic examination of acute and mid-term CMR outcomes, and in the use of a clinically-recapitulative mouse model of MI. The study has a number of limitations. It did not assess the impact of iron on in-hospital outcomes, because events that occurred prior to discharge were not included in the analysis. Though the study inferred patient’s baseline iron status from the earliest sampling timepoint (presentation), it cannot be fully excluded that iron markers had already changed secondary to STEMI by this stage. The role of anaemia was not examined, because of the lack of haemoglobin measurements in this cohort. The 6-month CMR data must be interpreted with caution, because of the potential for bias towards patients with better functional capacity. Another limitation is that it cannot interpret the associations between iron markers sampled at later timepoints and CMR parameters, because these associations could reflect bidirectional effects. Another limitation is that the study did not have any means of assessing myocardial iron content in STEMI patients, because T1-mapping at this stage is most profoundly affected by myocardial injury, rather than myocardial iron content.

## AUTHORS CONTRIBUTION

S.L.-L. conceived project, secured funds, analysed data, and wrote the manuscript. M.V.A and S.N.K collected pre-clinical data. M.V.A, S.N.K and G.M measured iron markers in human serum. M.S and R.K collected clinical data. M.D.C analysed all clinical data. K.M.C and J.G.C reviewed and commented on the manuscript. All the authors reviewed the manuscript prior to submission.

## CONFLICT OF INTEREST

S.L.-L. reports receipt of previous research funding from Vifor Pharma, personal honoraria on a lecture from Pharmacosmos and consultancy fees from Disc Medicine and ScholarRock.

## FUNDING

S.L.-L, MVA and SNK were funded by a Medical Research Council Senior Research Fellowship awarded to S.L-L (MR/V009567/1/) and the British Heart Foundation Centre for Research Excellence (HSR00031 and RE/18/3/34214). JGC is supported by the British Heart Foundation Centre of Research Excellence (RE/18/6134217).

## DATA AVAILABILITY

Research data will be made available upon reasonable request to the corresponding author.

## ETHICAL APPROVAL

The trial protocol and amendments were approved by a NHS ethics committee in the UK (REC: 11/SC/0397), and the Health Research Authority.

## SUPPLEMENTAL FIGURES

**Supplemental figure 1.**
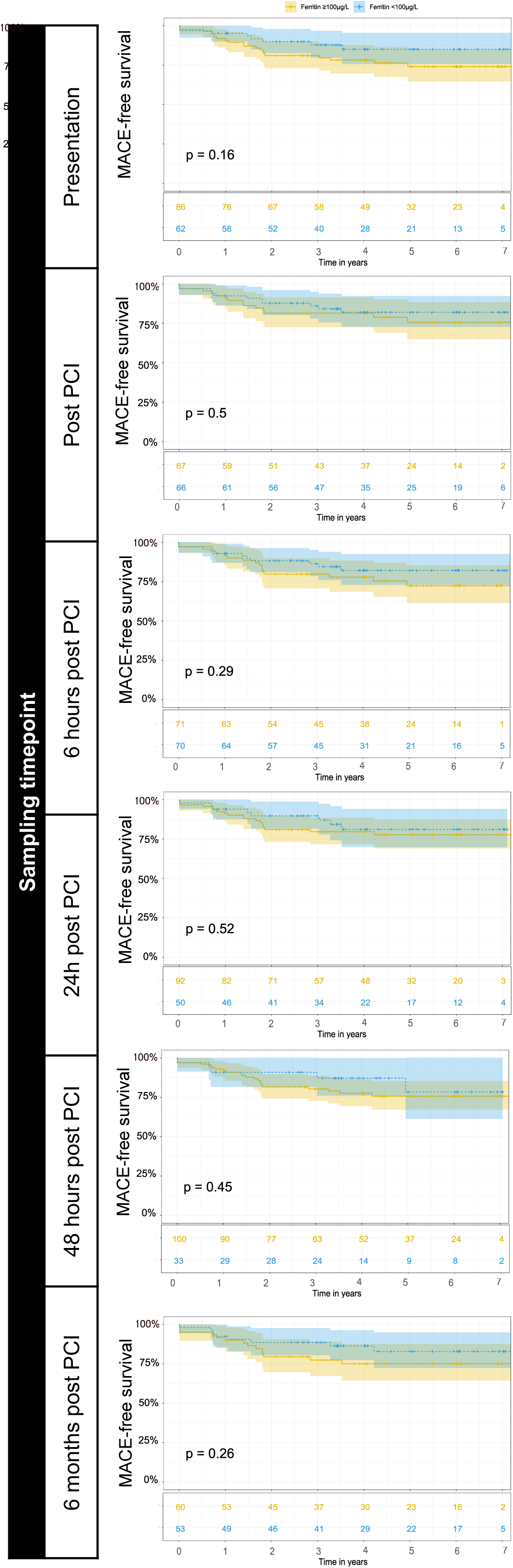
Hypoferritinemia is not associated with MACE-free survival after STEMI. Kaplan Meier survival plots showing percentage MACE-free survival. For each plot, patients were stratified according to whether they had ferritin<100ug/L or not at presentation (top row), at post PCI (second row), 6 hours post PCI (third row), 24 hours post PCI (fourth row), 48 hours post PCI (fifth row) or at 6 month follow-up (bottom row). P values shown were calculated using long rank test.

**Supplemental figure 2.**
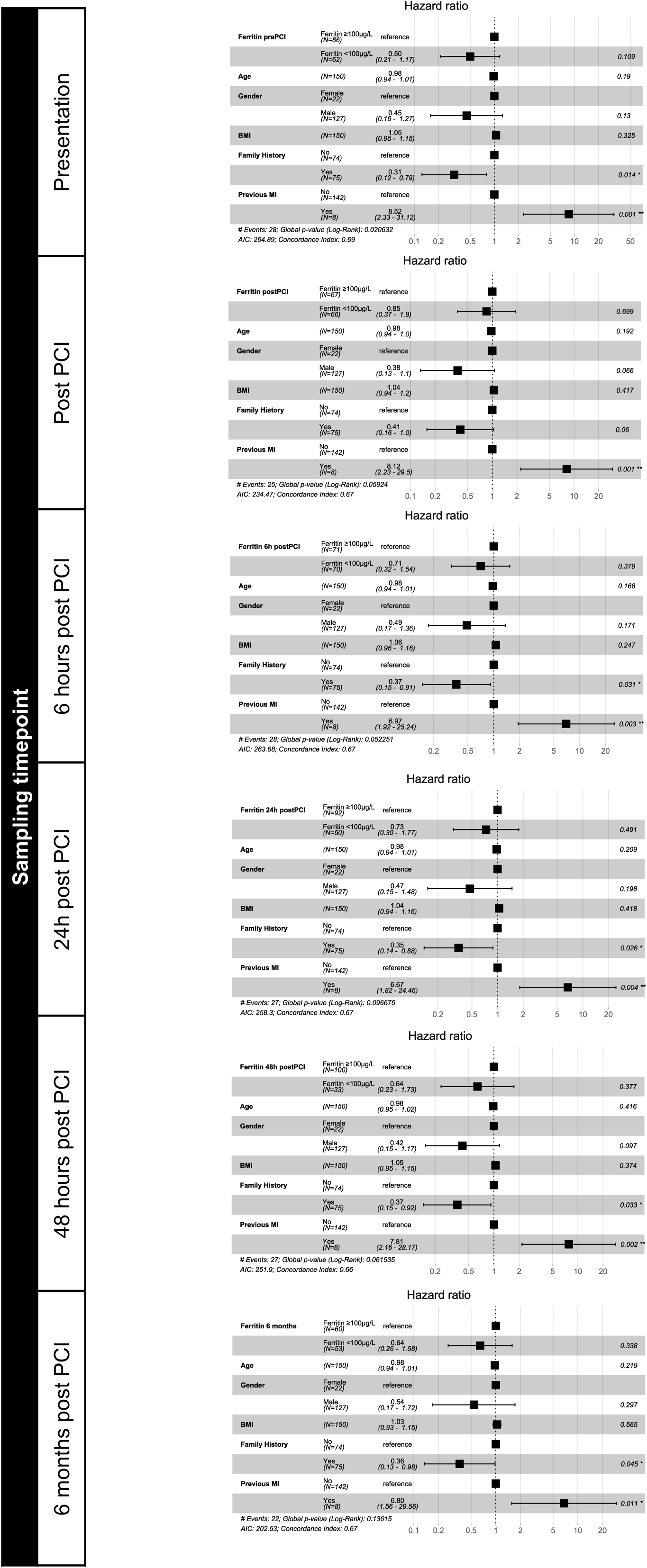
Hypoferritinemia does not predict risk of MACE after STEMI. Cox regression analysis for risk of MACE, based on the most parsimonious model. Forest plots are shown for the presence of ID at baseline, defined as ferritin<100ug/L. Modelling was run separately for every iron sampling timepoint.

**Supplemental figure 3.**
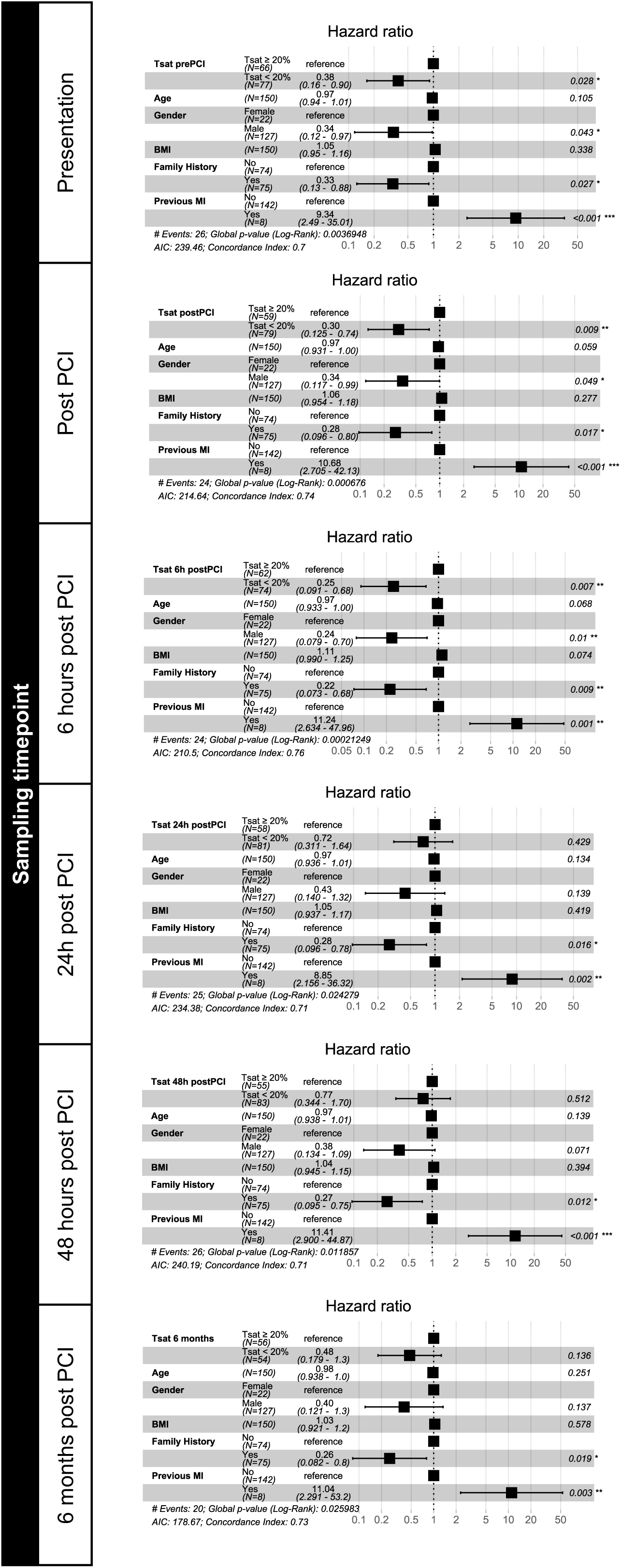
Complete forest plots depicting adjusted hazard ratios for MACE according to the presence of ID defined as Tsat<20%. Cox regression analysis for risk of MACE was based on the most parsimonious model. Modelling was performed separately for each sampling timepoint

**Supplemental figure 4.**
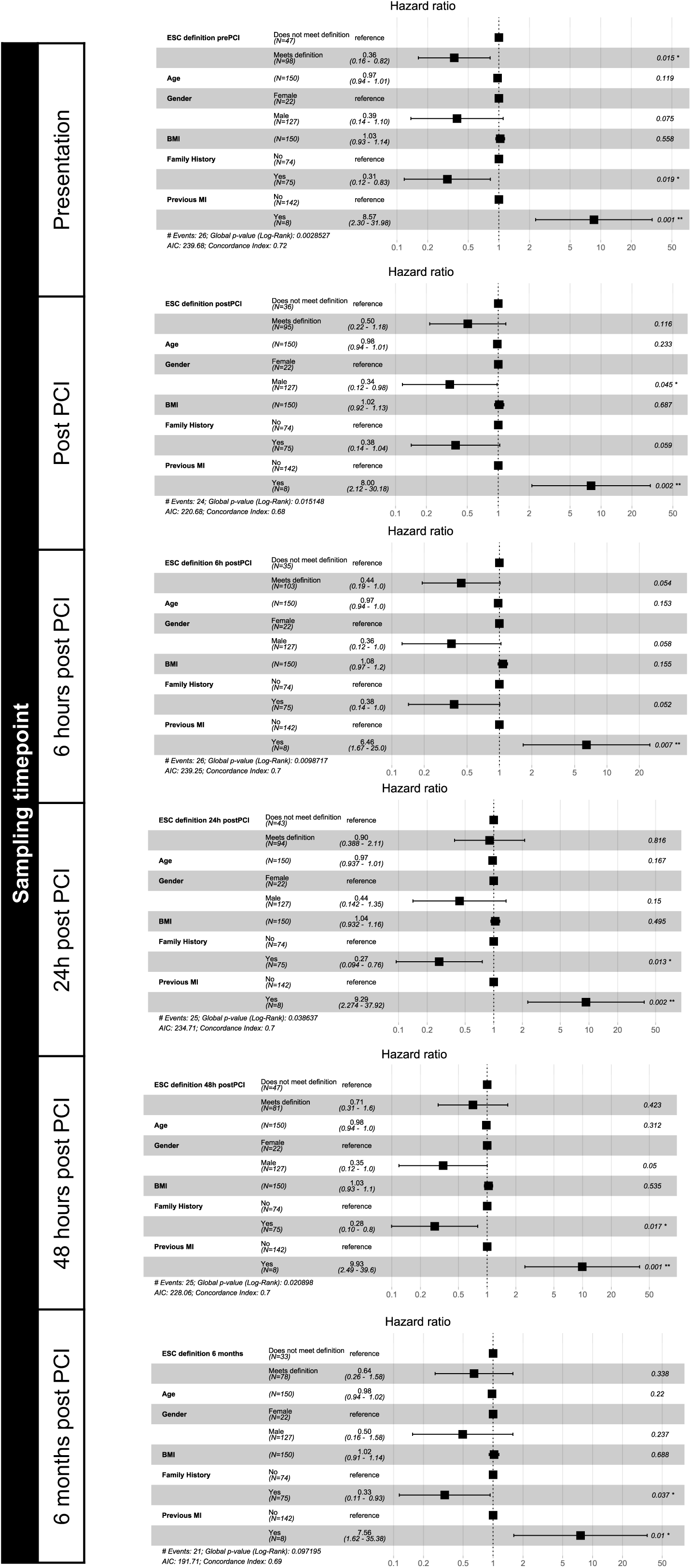
Complete forest plots depicting adjusted hazard ratios for MACE according to the presence of ID as defined by ESC. Cox regression analysis for risk of MACE was based on the most parsimonious model. Modelling was performed separately for each sampling timepoint

**Supplemental figure 5.**
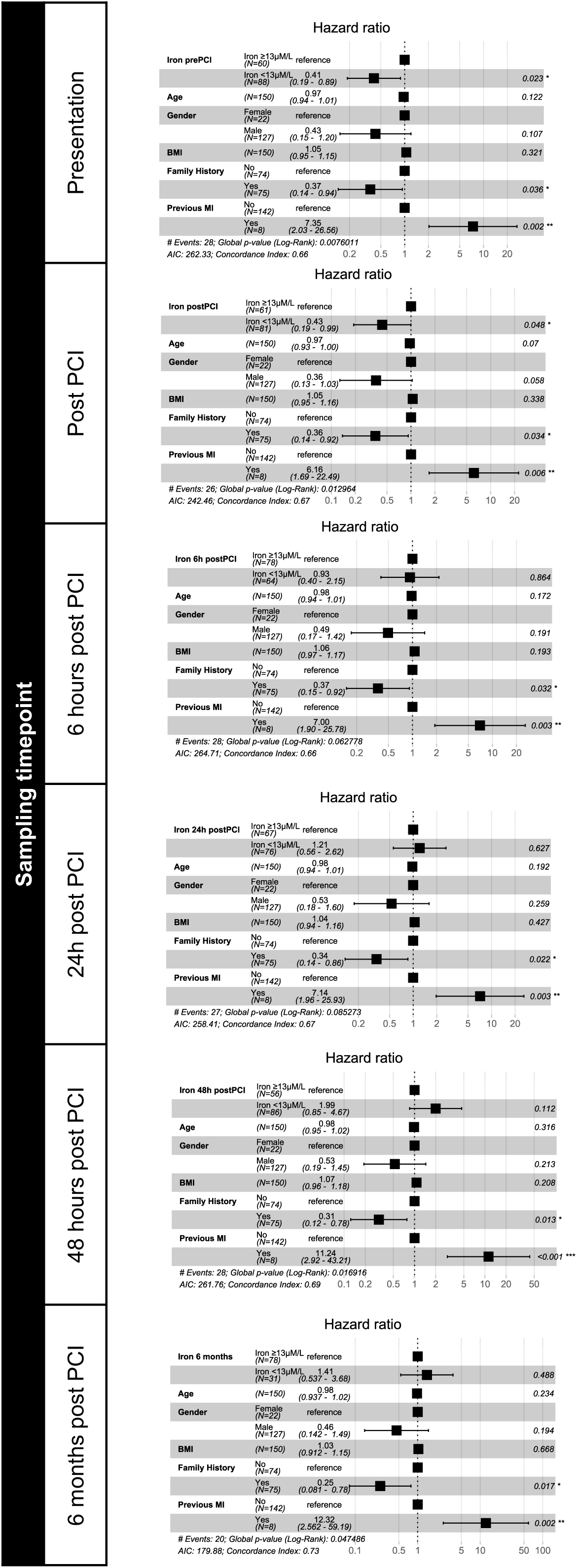
Complete forest plots depicting adjusted hazard ratios for MACE according to the presence of ID defined as serum iron<13uM. Cox regression analysis for risk of MACE was based on the most parsimonious model. Modelling was performed separately for each sampling timepoint

**Supplemental figure 6.**
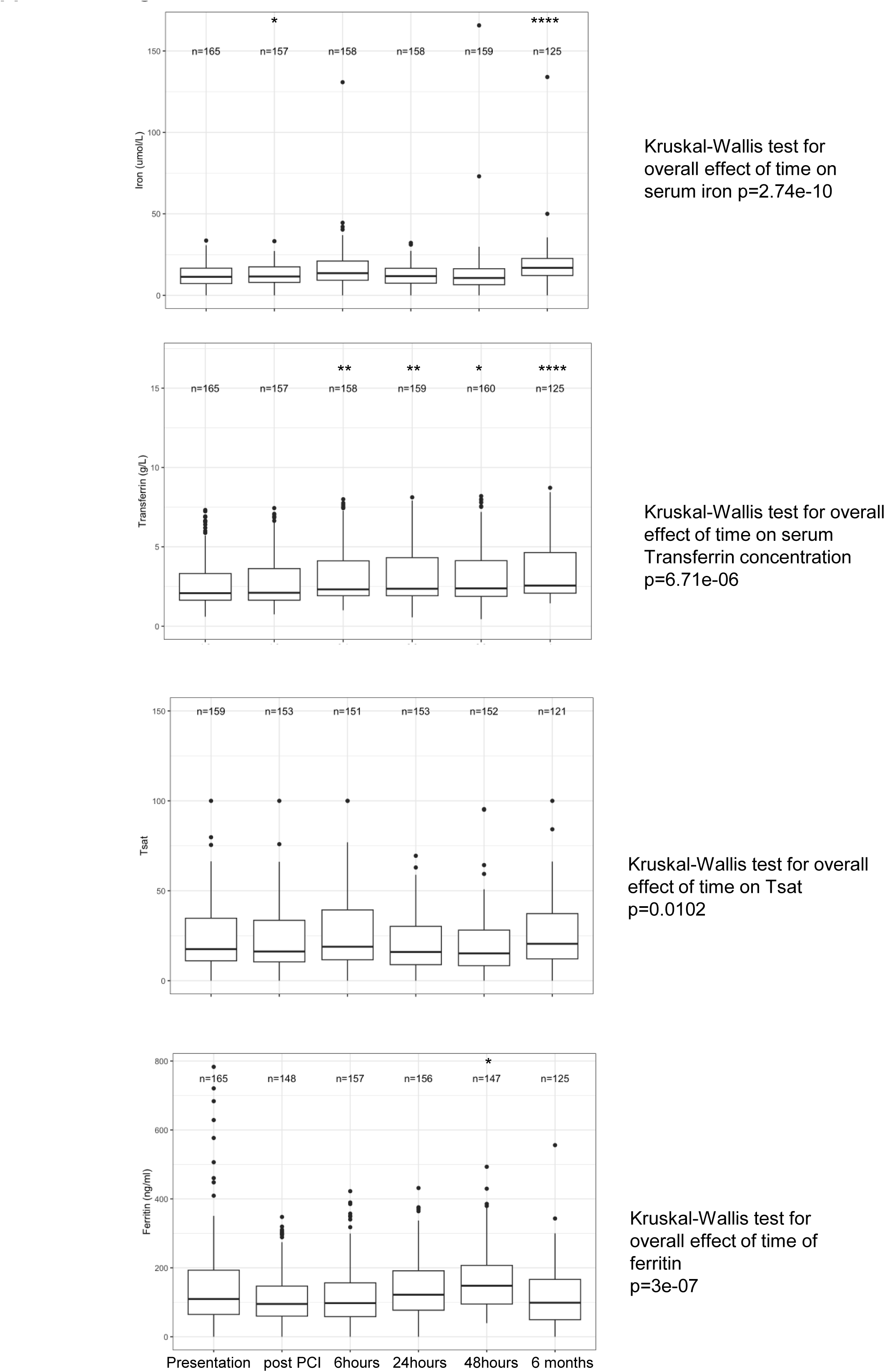
Serum iron markers change acutely after STEMI. A. Changes in serum iron concentration B. Changes in serum transferrin concentration C. Changes in Tsat D. Changes in test ferritin concentration Kuskall-Wallis test was used to test the overall effect of time on each marker. Post-hoc analysis was used to compare each timepoint to baseline (presentation). *p<0.05, **p<0.01, ***p<0.001 relative to presentation.

**Supplemental table 1.**
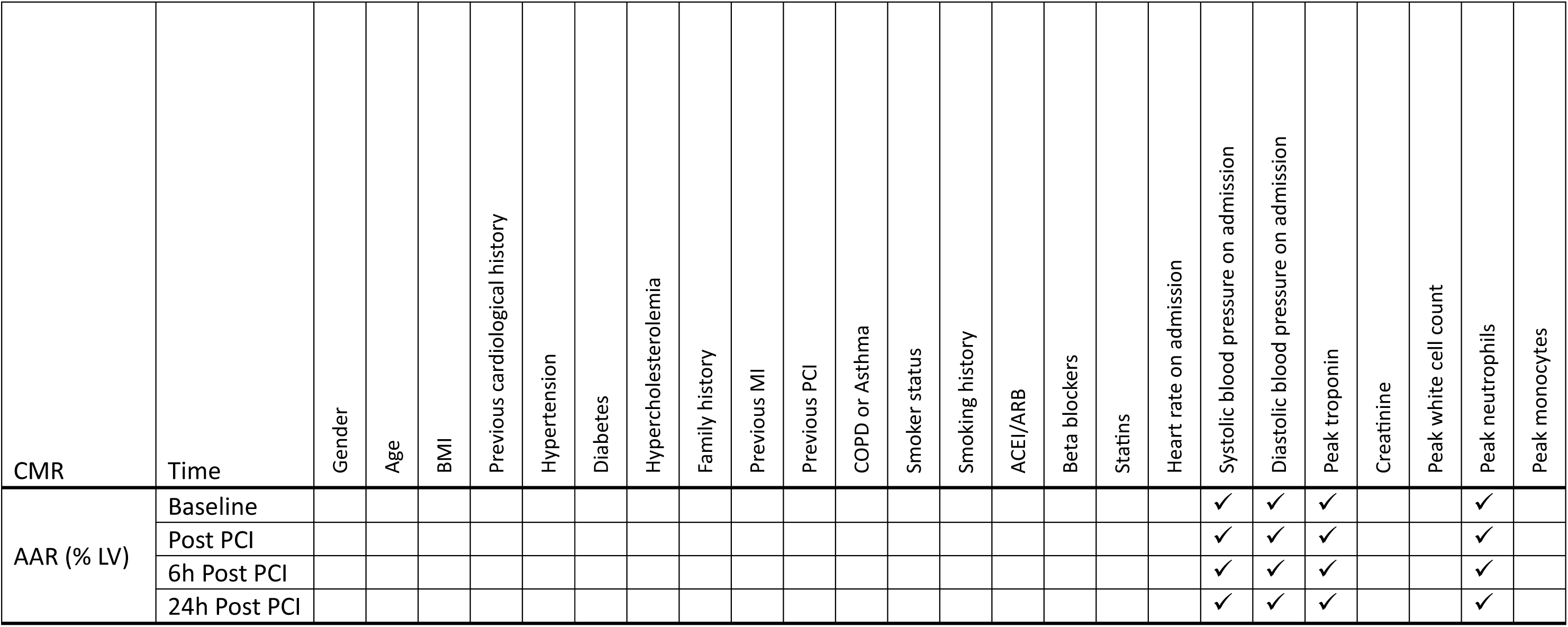
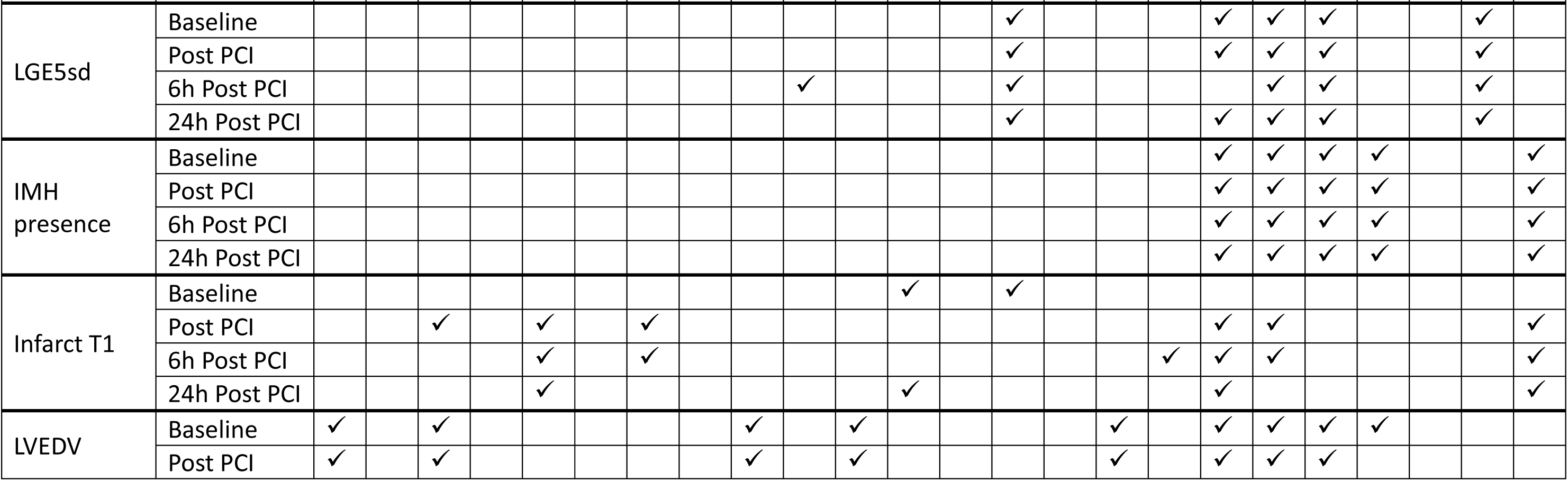
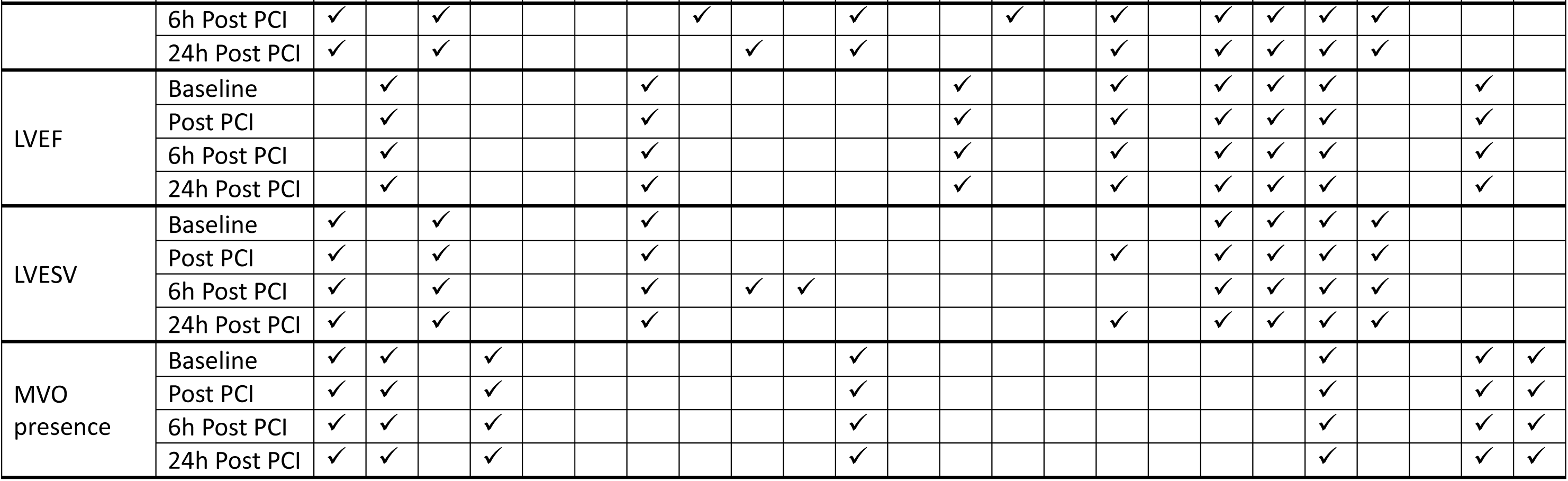
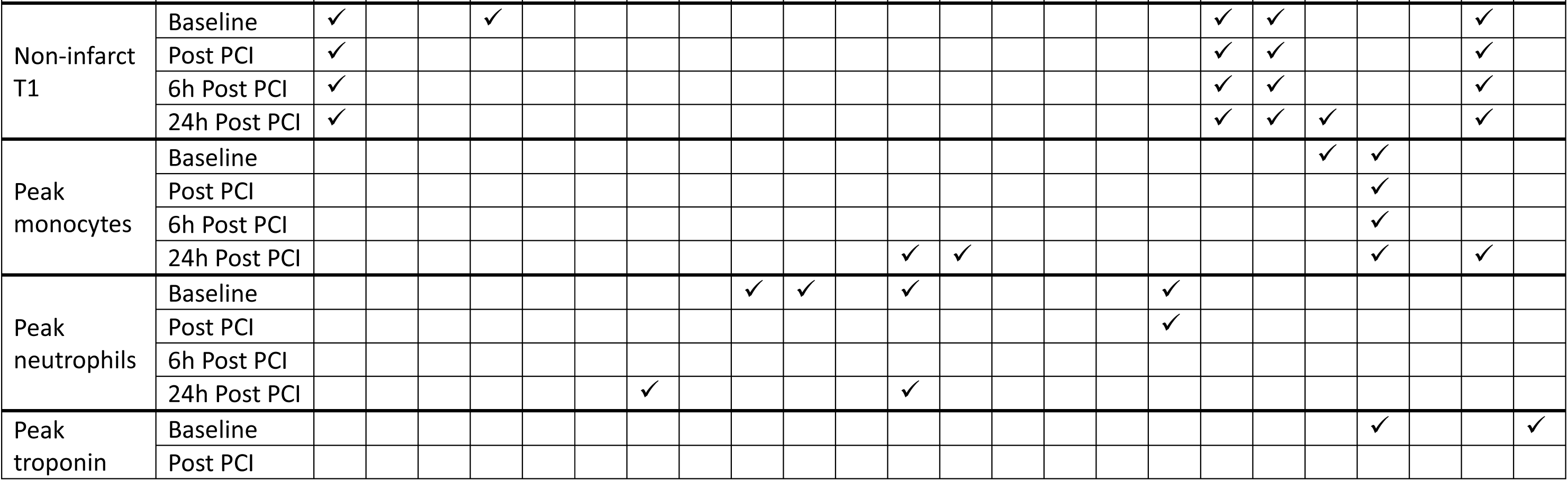
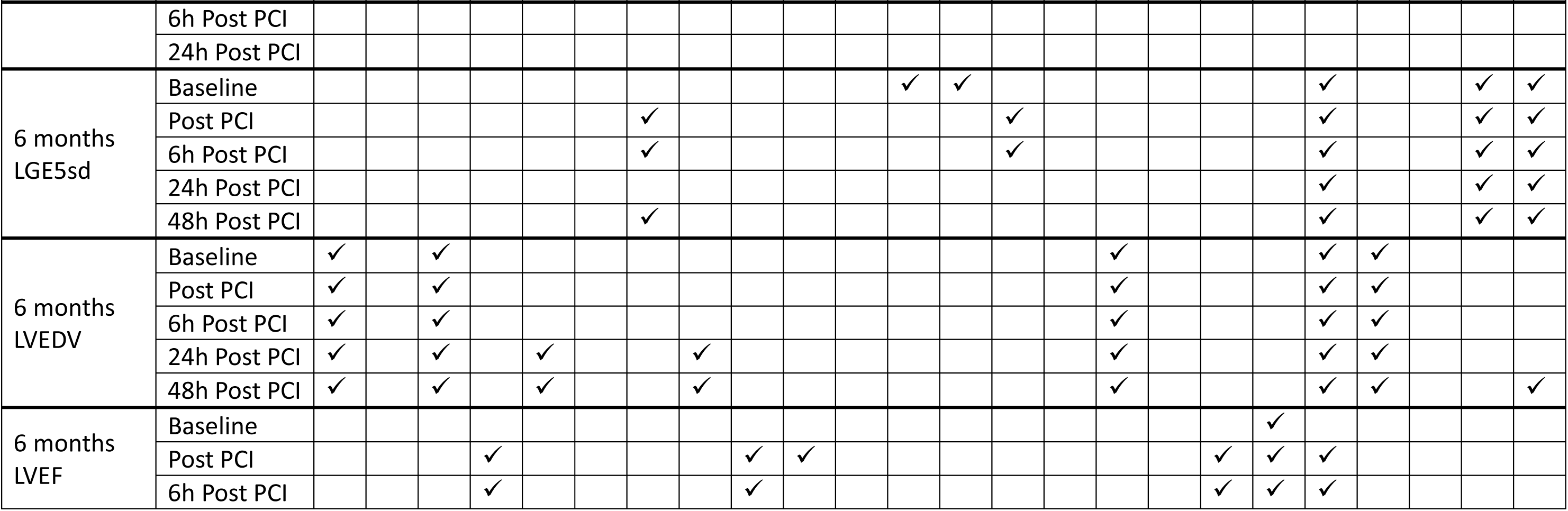
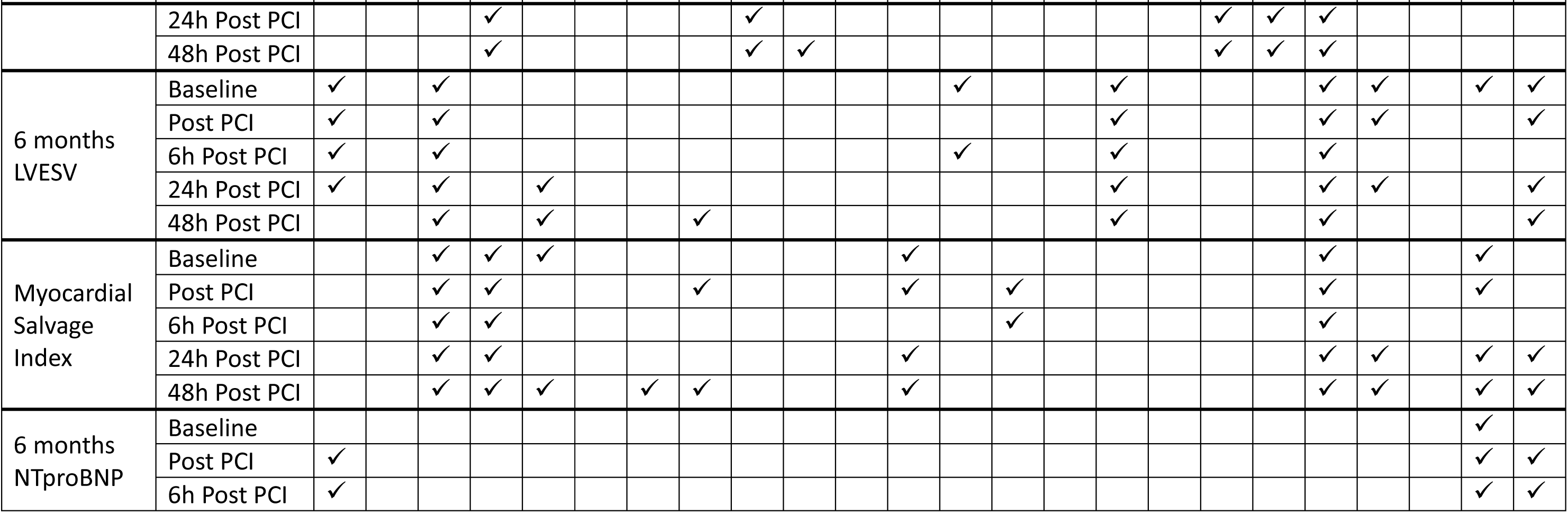
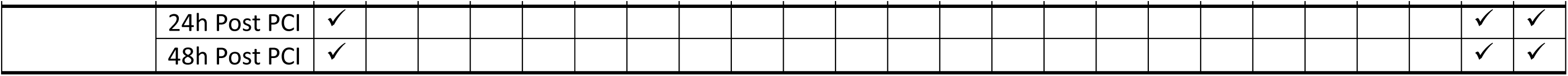
Details of modelling co-variates.

## SUPPLEMENTAL METHODS

### Mouse study design

To minimise any potential confounding effects of age, all mice were provided their respective diet at 4±0.5 weeks of age, and underwent surgical intervention for MI at 11±1 weeks of age. Randomisation was carried out using the Rand() function in excel. No animals were excluded from analysis

### MI in mice

MI was induced by ligation of the left anterior ascending artery (LAD). Briefly, buprenorphine (buprenorphine hydrochloride; Vetergesic) was delivered as a 0.015 mg/mL solution via intraperitoneal injection at 20 min before the procedure to provide analgesia. Mice were then anesthetized and maintained under 2.5% isoflurane under assisted external ventilation through the insertion of an endotracheal tube (~200 strokes/min; stroke volume ~200 μL/min). A lateral thoracotomy was performed, and an 8-0 suture used to ligate in the region of the LAD: just below the tip of the left atria, passing towards the centre of the heart. The ribs and skin were closed using 7-0 sutures and the mouse allowed to recover in a heated chamber. SHAM surgery controls underwent an identical protocol with the exception of the LAD ligation step.

### Cardiac Function (Cine MRI) in mice

Mice were anaesthetized with 2% isofluorane in O2 and positioned supine in a purpose-built cradle. ECG electrodes were inserted into the forepaws, a respiration loop was taped across the chest and heart signals were monitored using a custom-built physiological motion gating device. The cradle was lowered into a vertical-bore, 11.7 T MR system with a 40 mm birdcage coil (Rapid Biomedical, Würzburg, Germany) and visualised using a Bruker console running Paravision 2.1.1. A stack of contiguous 1 mm thick true short-axis ECG and respiration-gated cine-FLASH images were acquired. The entire in vivo imaging protocol was performed in approximately 60 min. Image analysis was performed using ImageJ (NIH Image, Bethesda, MD). Left ventricular volumes and ejection fractions were calculated from the stack of cine images.

### Imaging myocardial reactive iron in mice

For imaging reactive iron in the heart, B6;FVB-Ptprca Tg(CAG-luc,-GFP)L2G85Chco Thy1a/J mice (JAX strain #025854) were used as our previous work (14). These mice harbour a luciferase luminescence reporter transgene. When injected with 25nM of iron-caged luciferin (ICL-1), presence of intracellular reactive iron allows the conversion of ICL-1 into the luciferase substrate, resulting in luminescence (14). As described above, mice were randomised to a normoferric or hypoferric diet, and underwent MI and SHAM surgeries. Six hours after surgery, mice were injected intravenously with ICL-1, hearts rapidly excised under anaesthesia and placed in an IVIS LUMINA system. They were imaged under auto settings, and luminescence collected for at least 5 minutes.

## Notes

### Competing Interest Statement

SLL reports receipt of previous research funding from Vifor Pharma, personal honoraria on a lecture from Pharmacosmos and consultancy fees from Disc Medicine and ScholarRock

### Funding Statement

S.L.-L, MVA, SNK and GM were funded by a Medical Research Council Senior Research Fellowship awarded to S.L-L (MR/V009567/1/) and the British Heart Foundation Centre for Research Excellence (HSR00031 and RE/18/3/34214). JGC is supported by the British Heart Foundation Centre of Research Excellence (RE/18/6134217).

### Author Declarations

The OxAMI study trial protocol and amendments were approved by a NHS ethics committee in the UK (REC: 11/SC/0397), and the Health Research Authority.

### Summary of Updates

One author, Goran Mohammad, has been added to the author list.

